# Expression and polymorphism of genes in gallstones

**DOI:** 10.1101/2025.08.13.25333550

**Authors:** Wu Yifeng, Ma Qiang, Xue Rongquan, Sun Yuetong

**Affiliations:** Department of Hepatobiliary, Pancreatic and Spleen Surgery, Inner Mongolia Bayannur Hospital, Bayannur 015000, China; Department of Hepatobiliary, Pancreatic and Spleen Surgery, First Affiliated Hospital of Baotou Medical College, Baotou 014000, China; Department of Hepatobiliary, Pancreatic and Spleen Surgery, Inner Mongolia Autonomous Region People’s Hospital, Hohhot 010017, China; Inner Mongolia Autonomous Region People’s Hospital, Hohhot 010017, China

**Keywords:** cholesterol gallstones, KLF14, SR-B1, gene, polymorphism, dyslipidemia

## Abstract

Through the method of clinical case control study, to explore the expression and genetic polymorphism of KLF14 gene (rs4731702 and rs972283) and SR-B1 gene (rs5888 and rs838880) in patients with cholesterol gallstones, to analyze the correlation between KLF14 gene and SR-B1 gene, and to study whether the gene locus of KLF14 gene and SR-B1 gene are different and correlated with blood lipid indexes, gender, nationality and environmental factors in patients with cholesterol gallstones.

**Summary:** *Objective:* through the method of clinical case control study, to explore the expression and genetic polymorphism of KLF14 gene (rs4731702 and rs972283) and SR-B1 gene (rs5888 and rs838880) in patients with cholesterol gallstones, to analyze the correlation between KLF14 gen e and SR-B1 gene, and to study whether the gene locus of KLF14 gene and SR-B1 gene are different and correlated with blood lipid indexes, gender, nationality and environmental factors in patients with chole sterol gallstones.

*Methods:* A total of 200 patients undergoing elective laparoscopic cholecystectomy in our department were randomly selected, and the cholesterol gallstones group (100 cases) was taken as the case group according to the inclusion criteria. A total of 100 cases in the non-gallstone group were used as the control group. The DNA from peripheral venous blood of all subjects was extracted, and SNP typing was performed on the KLF14 gene (rs4731702, rs972283) and SR-B1 gene (rs5888, rs838880) in the case group and the control group by direct sequencing. To compare the genotypic and allelic frequencies of the KLF14 gene (rs4731702, rs972283) and SR-B1 gene (rs5888, rs838880) between the two groups, to explore whether there is an association between them, and to analyze the association between the locus of the two genes and blood lipid indicators as well as gender, ethnicity, and environmental factors in patients with gallbladder cholesterol gallstones.

*Results:* 1. Serum levels of triglyceride (TG), total cholesterol (TCH), low density lipoprotein (LDL), and apolipoprotein B(Apo-B) in the case group were significantly higher than those in the control group; The levels of high density lipoprotein (HDL) and apolipoprotein AI(Apo-AI) were significantly lower than those in the control group (P < 0.05). 2. The distribution differences of allele frequency and genotype frequency at rs4731702 of 2.KLF14 gene in case group and control group were statistically significant (P < 0.05); The difference in frequency distribution of alleles C and T was statistically significant (P < 0.05), and the frequency of allele C was higher than that of allele T, suggesting that allele C might increase the risk of cholesterol gallstones (OR = 1.547). In the case group, TG levels in CC genotype at rs4731702 of KLF14 gene were higher than those in CT and TT genotypes (P < 0.05). HDL, Apo-AI and Apo-AI/Apo-B levels in the CC genotype were lower than those in the CT and TT genotypes (P < 0.05). There were no significant differences in TCH, LDL or Apo-B (P > 0.05). 3. The distribution differences of allele frequency and genotype frequency at rs5888 locus of SR-B1 gene in case group and control group were statistically significant (P < 0.05); There were significant differences in frequency distributions of alleles C and T between the two groups (P < 0.05). The CC genotype at rs5888 of SR-B1 gene in the case group was significantly higher than that of CT+TT genotype (P < 0.0 5), suggesting that the mutant CC genotype might be a risk factor for cholesterol gallstones (OR = 2.279). The frequency of the C allele was higher than that of the T allele, indicating that the C allele might increase the risk of cholesterol gallstones (OR = 1.898). The gene frequency of the TT type gene in the case group was significantly lower than that of the control group, and HDL-C of the TT type gene was significantly higher than those of CC and CT type, while TG and TC H contents were significantly lower than those of CC and CT type (P < 0.05). LDL, Apo-AI, Apo-B and Apo-AI/ Apo-B had no statistically significant difference (P > 0.05). 4. The distribution differences of allele frequency and genotype frequency of rs838880 of SR-B1 gene in case group and control group were statistically significant (P < 0.05). The number of CC genotype at rs838880 of SR-B1 gene in case group was significantly lower than that in control group, which indicated that genotype CC at rs838880 of SR-B1 gene was the protection genotype. The TCH of patients with TT genotype in the case group was significantly higher than those of patients with TC and CC genotype, and the difference was statistically significant (P < 0.05). HDL of patients with CC type gene was significantly higher than those of patients with TC and TT type in this group (P < 0.05). This indicates that the C allele may be associated with an increase in HDL-C in blood. There were no significant differences in TG, LDL, Apo-AI and Apo-B (P > 0.05), and no significant differences in the distribution of allele and genotype frequencies of rs972283 in 5.KLF14 gene between the case group and the control group (P > 0.05). The TG and TCH levels in the GG genotype at rs972283 of the KLF14 gene in the case group were higher than those in the GA and AA genotypes (P < 0.05). HD L, Apo-AI and Apo-AI/ Apo-B levels in the GG genotype were lower than those in the GA and AA genotypes, and the differences were statistically significant (P < 0.05). There was no significant difference between LDL and Apo-B (P > 0.05). 6. Comparison of the genotypes CC, CT and TT of the KLF14 gene rs4731702 in the case group revealed that there was no significant difference among ethnic groups (P>0. 05), while the gender difference was statistically significant (P<0.05). 7.BMI, family history of cholesterol gallstones and exercise habit were the influencing factors of cholesterol gallstones. 8. Patients with BMI > 26 and genotype (KLF14 rs4731702) CC were more likely to develop cholesterol gallstones (OR=16.379) than BMI < 26. Patients with a family history of gallstones with genotype (KLF14 rs4731702)CC had a 6.689-fold increased risk (OR=6.689) of developing CC compared with patients without a family history of gallstones with genotype (KL F14 RS 473702) (P < 0.05). There was no statistical significance (P > 0.05) in the exercise habits of patients with genotype (KLF14 rs4731 702)CC.

*Conclusion:* 1. The single nucleotide polymorphism (SNP) at rs4731702 of 1.KLF14 gene may have gender distribution differences and be related to the susceptibility to cholesterol gallstones, and the carrying of allele C may increase the risk of cholesterol gallstones. 2. 2. The genotype CC of rs5888 in SR-B1 gene may be the risk genotype for the development of cholesterol gallstones. 3. genotype CC of sr-b1 gene rs 838880 may be the protective genotype for cholesterol gallstones, and allele c may be related to the increase of HDL-C in blood. 4. snps of 4.KLF14 and SR-B1 have significant correlation with dyslipidemia and lipoprotein metabolism. 5. Patients with cholesterol gallstones have different degrees of lipid and lipoprotein metabolism abnormalities. 6. The increased 6.BMI and family history of cholesterol gallstones are the risk factors for cholesterol gallstones. Exercise habit is the protective factor of cholesterol gallstones. 7. The interaction between genetic factors and environmental factors promotes the occurrence of cholesterol gallstones.

---

Gallstone disease is currently one of the most common digestive system diseases, and epidemiological studies have shown that the prevalence of gallstones is still increasing year by year^[1]^. Cholesterol gallstones account for more than 70% of all gallstones and the major exogenous risk factors include high-calorie diet, obesity, lack of exercise, pregnancy, estrogen, advanced age, diabetes and dyslipidemia^[2, 3]^. In recent years, researchers have explored the causes of cholesterol gallstones in the gallbladder by a variety of ways and achieved some progress in many research fields, but the underlying cause is still inconclusive. At present, it is generally believed that abnormal blood lipoprotein and lipid metabolism are one of the main factors that lead to changes in the physicochemical properties of bile and its transformation to cholesterol gallstones^[4]^.

Hilongfu, etc^[5]^Our study also confirmed that dyslipidemia in patients with gallbladder cholesterol gallstones was characterized by significantly increased levels of serum total cholesterol (TC), low density lipoprotein cholesterol (LDL-C), apolipoprotein B(Apo-B), and triglycerides (TG), and significantly decreased high density lipoprotein cholesterol (HDL-C) and apolipoprotein AI(Apo-AI).

With the development of molecular biology and the continuous improvement of the whole genome sequencing, people have gradually deepened the study of cholesterol gallstones in the gene field. Epidemiological and familial studies have found that genetic factors account for about 25% of the total risk of gallbladder stones^[6]^. At the same time, studies have shown that more and more susceptible genes and their polymorphisms are involved in the process of lipid metabolism, and these mutation sites have also been confirmed to be closely related to the occurrence and development of cholesterol gallstones^[7]^.

Kruppel-like factors (KLF) family is a family of zinc finger transcription factors that can bind to GC-rich sequences, including 17 identified members, and plays an important role in the regulation of cell metabolism, proliferation, and differentiation.^[8]^. KLF14, as a member of the KLF family, also known as basic transcription factor binding protein 5, is widely expressed in mammalian tissues, including muscle, brain, heart, fat, and liver. The human KLF14 gene, located on chromosome 7 (7q32.3), is 1059bp in length and has a molecular weight of 33124Da. KLF14 regulates gene expression involved in many important physiological and pathological processes in organisms by binding to cis-regulatory sites rich in GT/GC in promoters or enhancers of target genes^[9]^.

SR-B1 is the only cell surface high-density lipoprotein (HDL) receptor found so far. It causes cholesterol to pass through the liver and into bile in the form of plasma HDL through selective uptake of cholesterol and cholesterol esters. A study outside China has shown that for mice with over-expression of SR-BI gene in the liver, plasma HDL cholesterol is ingested by the liver and enters the bile through SR-BI, thus increasing the cholesterol concentration in bile [10]. By contrast, in SR-B1-deficient mice, plasma cholesterol levels are elevated, and secretion of bile cholesterol is decreased [11]. SR-B1 gene is located on chromosome 12, and 12 exons have been identified so far. Synonymous single nucleotide polymorphisms (rs5888, T[ thymine] and C[ cytosine]) located in exon 8 have been shown to be associated with the expression level and function of SR-B1[12]. The minor allele T can cause changes in the secondary structure of mRNA, resulting in decreased expression and function of SR-B1. A larger number of studies have shown that rs5888 is associated with blood lipids and coronary heart disease risk in different populations [13–18]. A study outside China has shown that the rs5888 polymorphism of SR-B1 gene is associated with higher HDL cholesterol and lower triglyceride levels in non-Asian men. We also observed an association of the rs838880-C allele of SR-B1 with increases in plasma TC[19] and HDL cholesterol [20].

Genome-wide association studies (GWAS) showed that KLF14 was the main trans-regulator of fat gene expression, affecting the concentration or size of low-density lipoprotein (LDL), high-density lipoprotein (HDL) and very low-density lipoprotein (VLDL). At the same time, GWAS also confirmed the close relationship between the polymorphic sites of KLF14 gene and diseases related to lipid metabolism abnormalities, such as metabolic syndrome and atherosclerosis.^[21–23]^. A separate study has shown that single nucleotide polymorphisms (SNPs) in the KLF14 gene at the rs4731702 and rs972283 loci are associated with lower HDL-C levels and lower ApoA-I/ApoB ratios.^[24]^. In recent years, relevant research results have proved that KLF14 affects blood lipid metabolism by regulating lipid metabolism pathways, which may increase the risk of cholesterol gallstones.

At present, clinical studies on KLF14 gene and SR-B1 gene and their polymorphism in china have focused on lipid metabolism diseases such as hypertension and type 2 diabetes, and the correlation study with gallbladder cholesterol gallstones is rare. Therefore, in this study, direct sequencing was used to detect SNP typing at the loci of KLF14 (rs4731702, rs972283) and SR-B1 (rs5888, rs838880) genes in patients of the case group and the control group, aiming to explore the relationship between the polymorphisms of KLF14 and SR-B1 genes and the levels of cholesterol gallstones, blood lipids and lipoproteins in the gall bladder at the gene level. This will be of great importance for further research on the genetic mechanism of cholesterol stone formation, as well as for early screening and individualized prevention. At the same time, it will improve our understanding of the genetic mechanism of cholesterol stone formation and may make it possible to individualized prevention and screening of high-risk populations for gallstones.

## Information and methods

### 1. Subjects

#### 1.1 Cholesterol gallstones group

A total of 100 patients who were hospitalized with gallstones and underwent laparoscopic cholecystectomy in the Department of Hepatobiliary, Pancreatic, and Spleen Surgery of our hospital from December 2020 to April 2021 due to “gallstones with chronic cholecystitis” were randomly selected, including 42 males and 58 females, with the average age of 51.25 12.23 years old and the average Body Mass Index (BMI) of 25.14 1.63 kg/m2.

Inclusion criteria of cases: patients confirmed to have gallbladder stones after elective laparoscopic cholecystectomy; Preoperative auxiliary examination revealed: gallstones; Preoperative biochemical test results were normal in all cases; There was no clinical manifestation of acute inflammation at the time of admission; The stones were classified as cholesterol stones by component analysis after operation. Postoperative pathological analysis of the gallbladder tissue revealed chronic inflammatory changes.

Exclusion criteria of cases: patients with acute cholecystitis or acute cholangitis within 1 to 2 months; History of chronic diseases such as coronary heart disease, diabetes, hypertension, hepatitis and serious diseases of important organs; Patients with purulent or white bile found during surgery; Postoperative pathological results of gallbladder tissues suggested acute inflammatory changes or malignant changes.

#### 1.2 Non-gallstone group

A total of 100 patients who were hospitalized and underwent laparoscopic cholecystectomy for gallstones due to “gallbladder polyps or gallbladder adenomyosis” in the Department of Hepatobiliary, Pancreatic and Spleen Surgery of our hospital from December 2020 to April 2021 were randomly selected, including 40 males and 60 females, with the average age of 52.32 13.15 years old and the average BMI of 24.42 1.03 kg/m2.

Criteria for case inclusion: patients confirmed as gallbladder polyps or adenomyosis after elective laparoscopic cholecystectomy; Preoperative auxiliary examination shows: gallbladder polyps or gallbladder adenomyosis; Preoperative biochemical test results were normal in all cases; There was no clinical manifestation of acute inflammation at the time of admission; There was no acute inflammatory change or malignant change in the pathological results of gallbladder tissues after operation.

Exclusion criteria of cases: patients with acute cholecystitis or acute cholangitis within 1 to 2 months; History of chronic diseases such as coronary heart disease, diabetes, hypertension, hepatitis and serious diseases of important organs; Patients with purulent or white bile found during surgery; Postoperative pathological results of gallbladder tissues suggested cholesterol polyps, acute inflammatory changes or malignant changes.

This study was approved by the Ethics Committee of our hospital and informed consent was obtained from the subjects.

### 2. research methods

#### 2.1 Experimental Steps

##### 2.1.1 General information collection

General data including the name, gender, age, ethnicity, height, weight, BMI, smoking and alcohol consumption history, exercise habit and family medical history of the patients were collected using our inpatient case system and face-to-face interviews.

##### 2.1.2 Blood sample collection

After overnight fasting with water for all subjects, the medical staff of hepatobiliary surgery in our hospital took two tubes of venous blood (about 3–4 mL each) in the fasting supine position of the enrolled patients on the next morning and collected them into 5ml disposable EDTA anticoagulation vacuum blood collection tube. After collection, the basic information and serial number of blood-like test subjects were recorded. The marked first tube was immediately sent to the Clinical Laboratory of our hospital for biochemical project examination and test. The second tube was immediately placed in the-80 C refrigerator for cryopreservation pending genetic polymorphism detection.

##### 2.1.3 Determination of blood lipid and lipoprotein

Serum TC, TG, HDL, LDL, Apo-AI, Apo-B and other indicators were detected by automatic biochemical analyzer in the Department of Clinical Laboratory of our hospital.

##### 2.1.4 blood DNA extraction

(1) Thawing of blood samples: The blood samples were transferred from the-80 C refrigerator to the-20 C refrigerator for storage for 24 hours and then transferred to the-4 C refrigerator again. After 24 hours, the samples were thawed at room temperature and the sample DNA was extracted after the samples were completely in the liquid state.

(2) DNA extraction of blood sample: in this experiment, the Ezup column type blood genomic DNA extraction kit was used. Steps: ① The thawed blood samples were added into a 1.5 ml centrifuge tube, followed by PBS Solution to obtain a total liquid volume of 200 μl, and mixed evenly. ② Add 20 μl Proteinase K solution into the above centrifuge tube, mix well, add 200 µl Buffer DL, shake and mix well, and then place in a water bath at 56°C for 10 min. ③ Add Buffer DL 200 µl into the above centrifuge tube, mix with shaking, and perform a water bath at 56°C for 10 min. After 10 min, the mixed solution becomes clear and transparent, turning into complete lysis. When the cell lysis is incomplete, the solution still shows a turbid state, and the water bath time should be appropriately extended. ④ 200 µl anhydrous ethanol was added into the centrifuge tube and mixed evenly. ⑤ The adsorption column was put into a collection tube, and the semitransparent fibrous suspended matter and solution were all added into the adsorption column with a pipette, and the mixture was allowed to stand for 2 min. Then the mixture was centrifuged using a centrifuge at 10000 rpm for 1 min at room temperature, and the waste liquid was discarded. ⑥ Place the adsorption column back into the collection tube, add 500 µl GW Solution to the adsorption column, and centrifuge at 10,000 rpm for 0.5min before discarding the waste liquid. ⑦ The adsorption column was put back into the collection tube again, and 700 µl Wash Solution was added into the adsorption column. The waste liquid was discarded after being centrifuged for 0.5min at 10,00 rpm. ⑧ Repeat the previous experimental step. ⑨ Once again, put the adsorption column back into the collection tube, and centrifuge at 12000 rpm for 2 min at room temperature to remove the residual Wash Solution. ⑩ The removed adsorption column was placed in a new 1.5 ml centrifuge tube. 50 L CeBuffer was added and allowed to stand for 3 min. After the column was centrifuged at 12,000 rpm for 2 min at room temperature, the DNA solution was collected. The extracted DNA was stored in a refrigerator at-20 C or subjected to the next experiment.

##### 2.1.5 SNP typing test

1. Direct sequencing: As the gold standard of SNP typing, direct sequencing has the advantages of intuition, high accuracy and the like. The method comprises the following steps of: designing amplification primers at two sides of an SNP site, expanding a fragment where a target SNP is located, and verifying the base condition of the SNP site through a result of one-generation sequencing.
2. primer design and synthesis: according to the national center for biotechnology information (NCBI) official website (http://www.ncbi.nlm.nlh.gov) and rs972283, Premier 5.0 software was used to design the primers, with the primer length of 18–30 bp, Tm value of 55–65℃, annealing temperature of about 60℃, and GC content of 40–70%. Non-specific amplification and primer dimer should be avoided. The length of PCR amplification products: the primers for site sequencing were about 150∼300 bp and 80–150 bp from the site, and the primers for explicit detection were about 150 bp from the upstream and downstream exons; The PCR product band of the target gene sequencing is not more than 1200 bp. Entrust shanghai shenggong bioengineering company to synthesize primer.

**Table 1:**
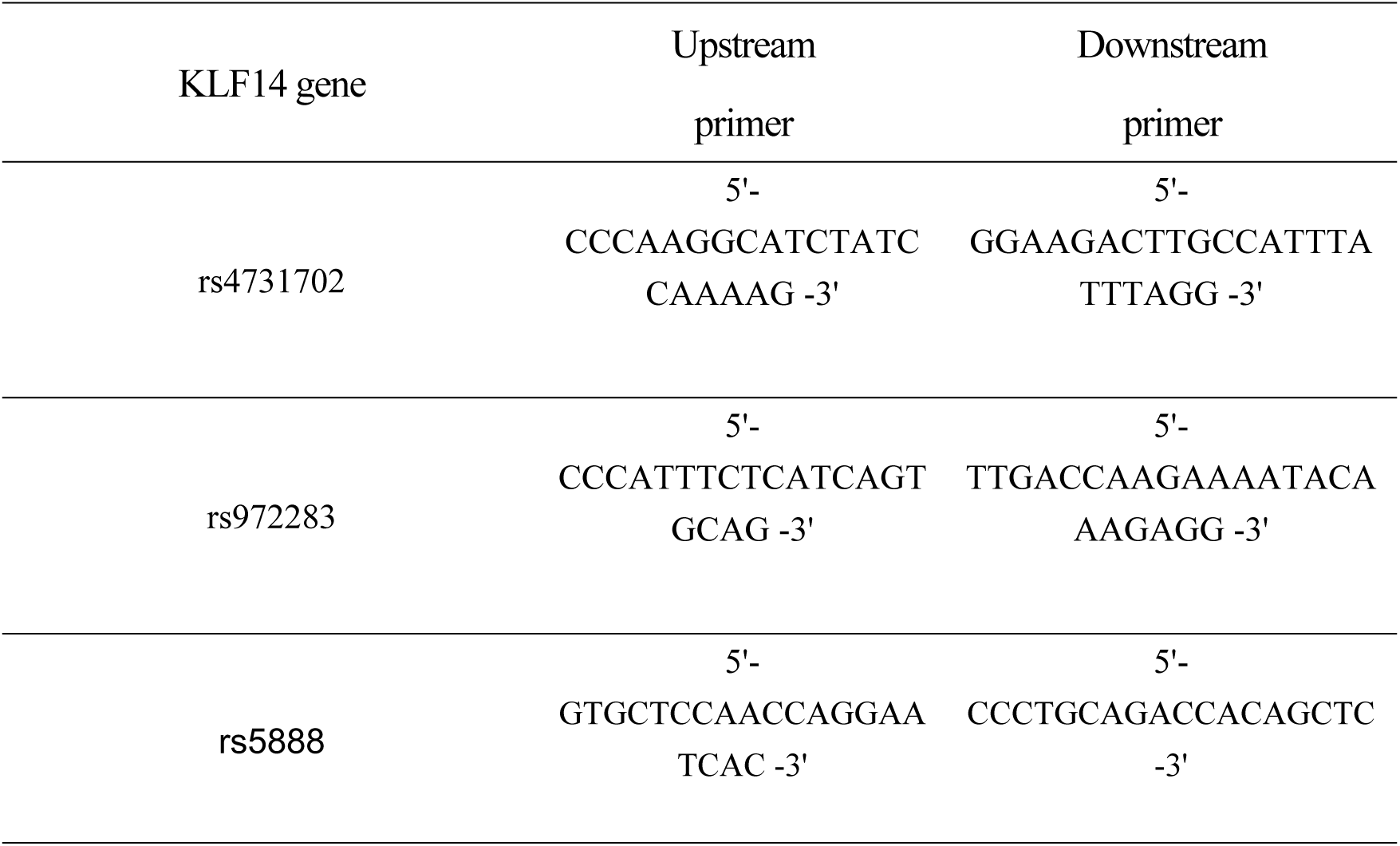
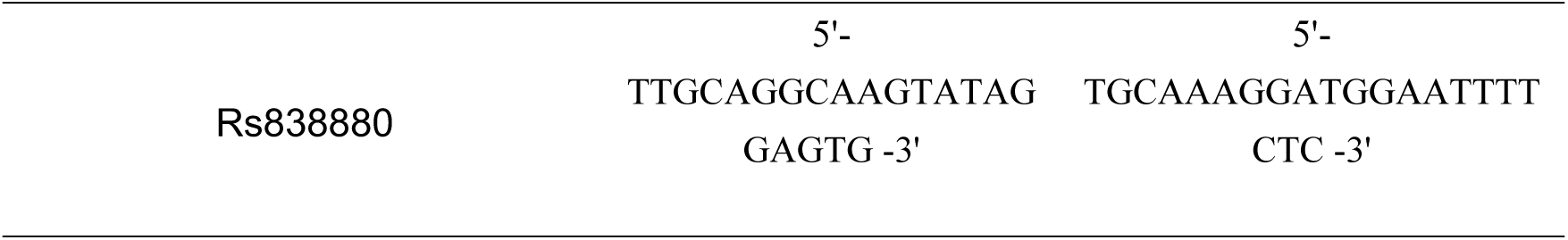
primer sequences for the KLF14 gene rs4731702 and rs972283.

##### 2.1.6 PCR detection

(1) Principle: Polymerase chain reaction (PCR) is a molecular biology technology used to amplify specific DNA fragments in vitro. The main proces of that PCR technology is that a short single-stranded DNA fragment synthesize by genetic material is used as a primer, the primer can be specifically combine with a specific region of the template DNA, and four dNTP are used as substrates, and the DNA fragment is formed by polymerization along the 3’ end of a double-stranded part formed by the primer and the template DNA with the participation of DNA polymerase to realize the in vitro amplification of the DNA. According to the DNA polymerase in 97 degrees above the high temperature can also maintain the stability of the characteristics, so in 94 ∼ 97 degrees in the temperature range of DNA to form a single chain, and then cooling to 50-55 degrees, until the right amount of primer binding to the template. Finally, the temperature was increased to 72 degrees, and the polymerase was polymerized again to form double-stranded DNA.

(2) PCR experimental steps: 1. DNA denaturation (90 C–96 C): under the action of heat, the double-stranded DNA template fractured hydrogen bonds to form single-stranded DNA. 2. Annealing (25 C–65 C): the system temperature was reduced, and the primers combined with DNA template to form local double chain. 3. Extension (70 C–75 C): The system temperature was adjusted to 72 C, and the activity of Taq enzyme was the strongest. Under the action of Taq enzyme, DNA strand complementary to the template was synthesized by extending from the 5 ′ end to 3 ′ end of dNTP using dNTP as the raw material.

(3) PCR reaction system:

**Table 2:** pcr reaction system.

| component | concentration | Volume (μl) |
| --- | --- | --- |
| Template DNA |  | one |
| Upstream primer | 10 μM | one |
| Downstream primer | 10 μM | one |
| DNTP mixture | 10 mM | one |
| Taq buffer containing MgCl<sub>2</sub> | 10X | 2.5 |
| taq enzyme | 5 U/μl | 0.2 |
| Sterilized deionized water |  | 25 |

(4) PCR reaction program

**Table 3:** pcr reaction procedure.

| serial number | step | Temperature (C) | Time (min) |
| --- | --- | --- | --- |
| one | Pre-denaturation | 95 | five |
| two | denature | 94 | 0.5 |
| three | anneal | 63 | 0.5 |
| four | extend | seventy-two | 0.5 |
| five | Repair extension | seventy-two | 10 |

Note: Denaturing, annealing, and extension (Steps 2-4 of Table 3) were cycled 40 times, followed by repair extension at 72 C for 10 min.

##### 2.1.7 purification of pcr product

In this experiment, the SanPrep column PCR product purification kit was used, and the procedures are shown in Table 4.

**Table 4:** purification of 6: PCR product.

| serial<br>number | Fuck |
| --- | --- |
| ① | Add the PCR reaction solution to a 1.5ml centrifuge tube, and add 5 times the volume of Buffer B3, and mix well. |
| ② | The mixed solutions were transferred to the adsorption column, and centrifuged at $8000\times g$ for 0.5min. The liquid in the collection tube was discarded, and the adsorption column was put into the same collection tube. |
| ③ | Then 500 $\mu$ l Wash Solution was added into the adsorption column, and the mixture was centrifuged at $9000\times g$ for 0.5min. The liquid in the collection tube was discarded, and the adsorption column was placed into the collection tube. |
| ④ | Repeat the previous step once. |
| ⑤ | The empty adsorption column and collection tube were placed in a centrifuge and centrifuged at $9000\times g$ for 60 seconds. |
| ⑥ | Add 30 $\mu$ L of Elution Buffer to the center of the adsorption membrane, allow to stand at room temperature for 2 minutes and centrifuge at $9000\times g$ for 60 seconds. The extracted DNA solution was stored at -20 c. |

##### 2.1.8 PCR product identification

The PCR products of case group and control group were taken and detected by agarose gel electrophoresis. DNA Marker of 100–10000 bp DNA molecular weight standard was used as the quality reference standard, as shown in Fig. 1. Lane M was the Marker Ladder, and lanes 1–4 were the PCR sample products. As a result, the target fragments of the amplification product at rs4731702 site of KLF14 gene were found to be 200–300 bp, and the electrophoresis target band length of the PCR product was 260bp, as shown in Fig. 2. The target fragment of the amplification product at rs972283 site of KLF14 gene was 200–300 bp, and the length of the electrophoresis target band of the PCR product was 295bp, as shown in Fig. 3. The target fragments of the amplification product at rs5888 site of SR-B1 gene were 200– 300 bp, and the electrophoresis target band length of the PCR product was 264bp, as shown in Fig. 4. The target fragment of the amplification product at rs838880 site of SR-B1 gene was 200–300 bp, and the length of the electrophoresis target band of the PCR product was 289bp, as shown in Fig. 5. It could be seen from the test results that the PCR products met the product length requirements of primer design, and sequencing experiment could be continued.

**Figure 1:**
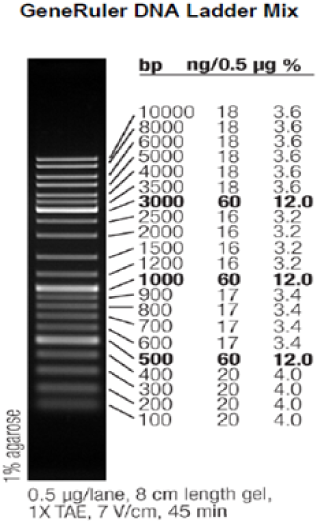
DNA marker (Sm0331)

**Figure 2:**
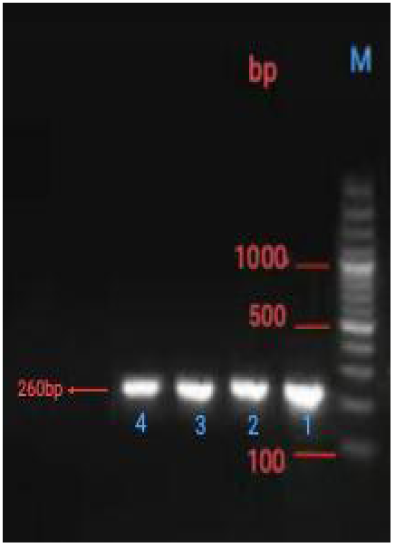
PCR chromatogram of RS 473702.

**Figure 3:**
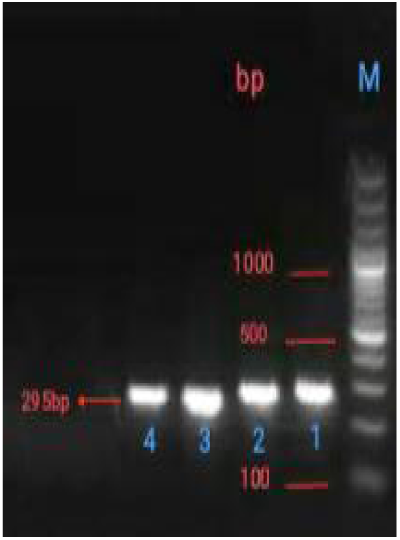
PCR chromatogram of RS 972283.

**Figure 4:**
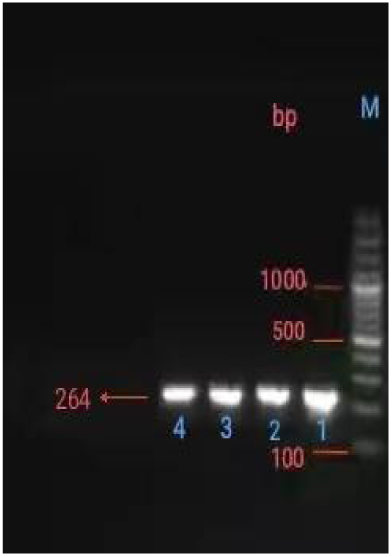
rs5888 pcr electrophoresis.

**figure 5:**
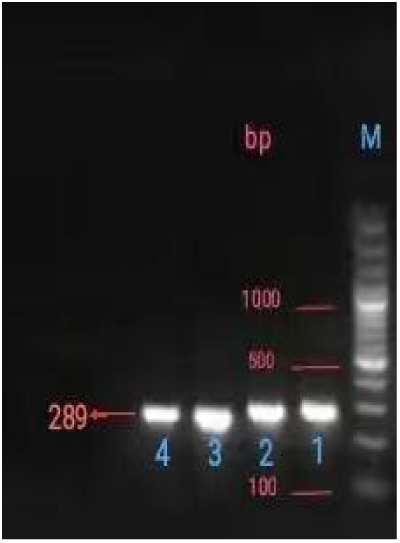
rs838880 pcr electrophoresis.

##### 2.1.9 PCR product sequencing

(1) Principle: The first generation sequencing technology (Sanger method) is based on the principle of dideoxy chain end termination method. According to the nucleotide starting at a fixed point and randomly terminating at a specific base, a series of nucleotides with four groups of A, T, C and G with different lengths are generated, which are then detected by electrophoresis on urea-denatured PAGE gel, thereby obtaining DNA sequences.

(2) PCR sequencing reaction system

**Table 5:**
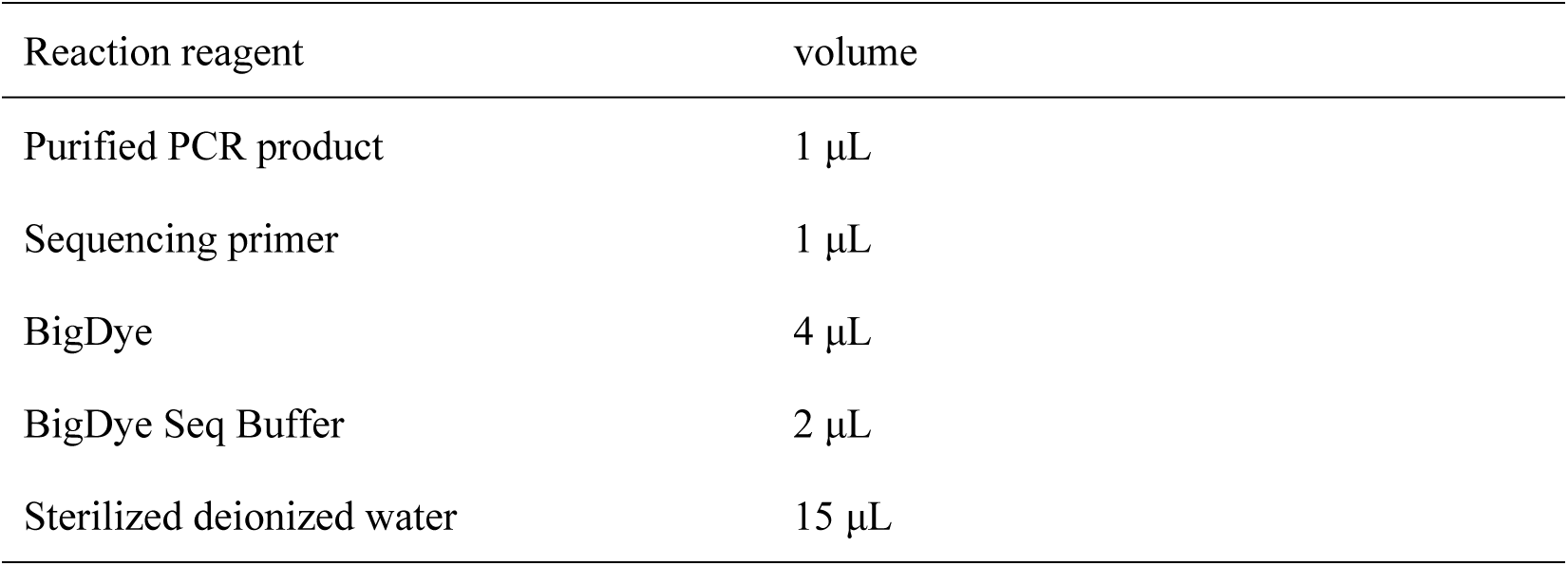
pcr sequencing reaction system.

| Reaction reagent | volume |
| --- | --- |
| Purified PCR product | 1 μL |
| Sequencing primer | 1 μL |
| BigDye | 4 μL |
| BigDye Seq Buffer | 2 μL |
| Sterilized deionized water | 15 μL |

(3) PCR sequencing reaction conditions

**Table 6:**
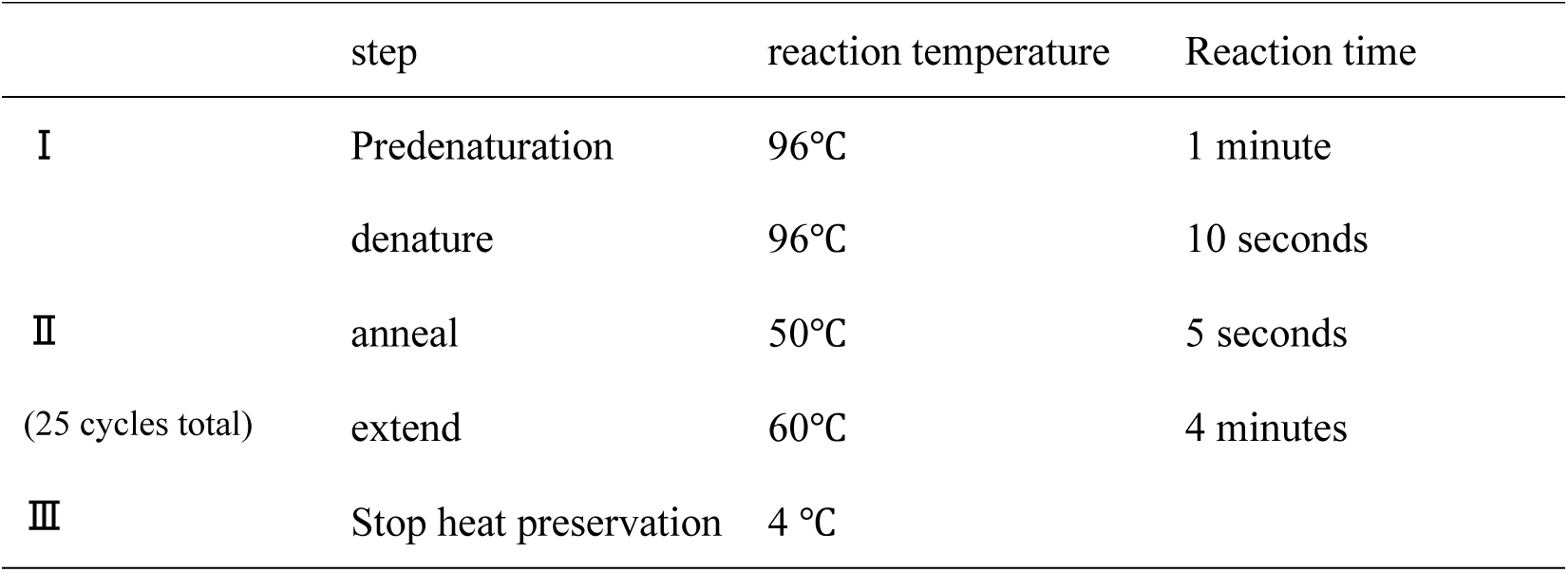
pcr sequencing reaction conditions.

|  | step | reaction temperature | Reaction time |
| --- | --- | --- | --- |
| I | Predenaturation | 96°C | 1 minute |
|  | denature | 96°C | 10 seconds |
| II | anneal | 50°C | 5 seconds |
| (25 cycles total) | extend | 60°C | 4 minutes |
| III | Stop heat preservation | 4 °C |  |

##### 2.1.10 purification of sequence products

**Table 7:**
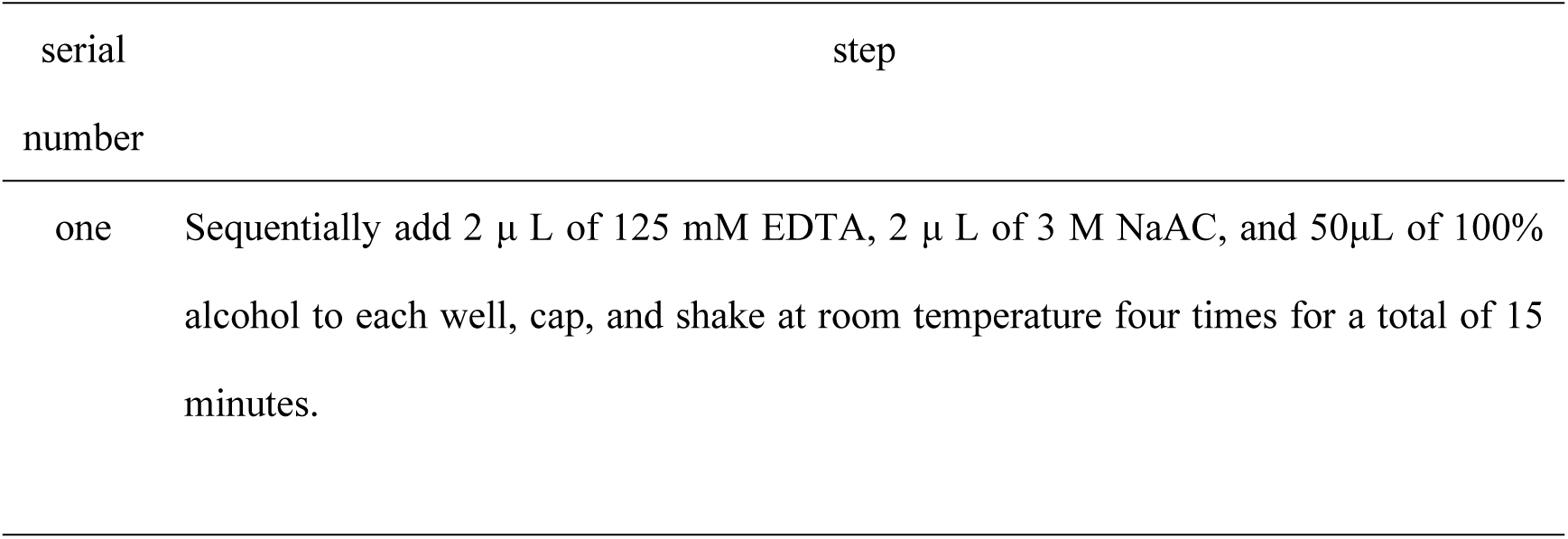
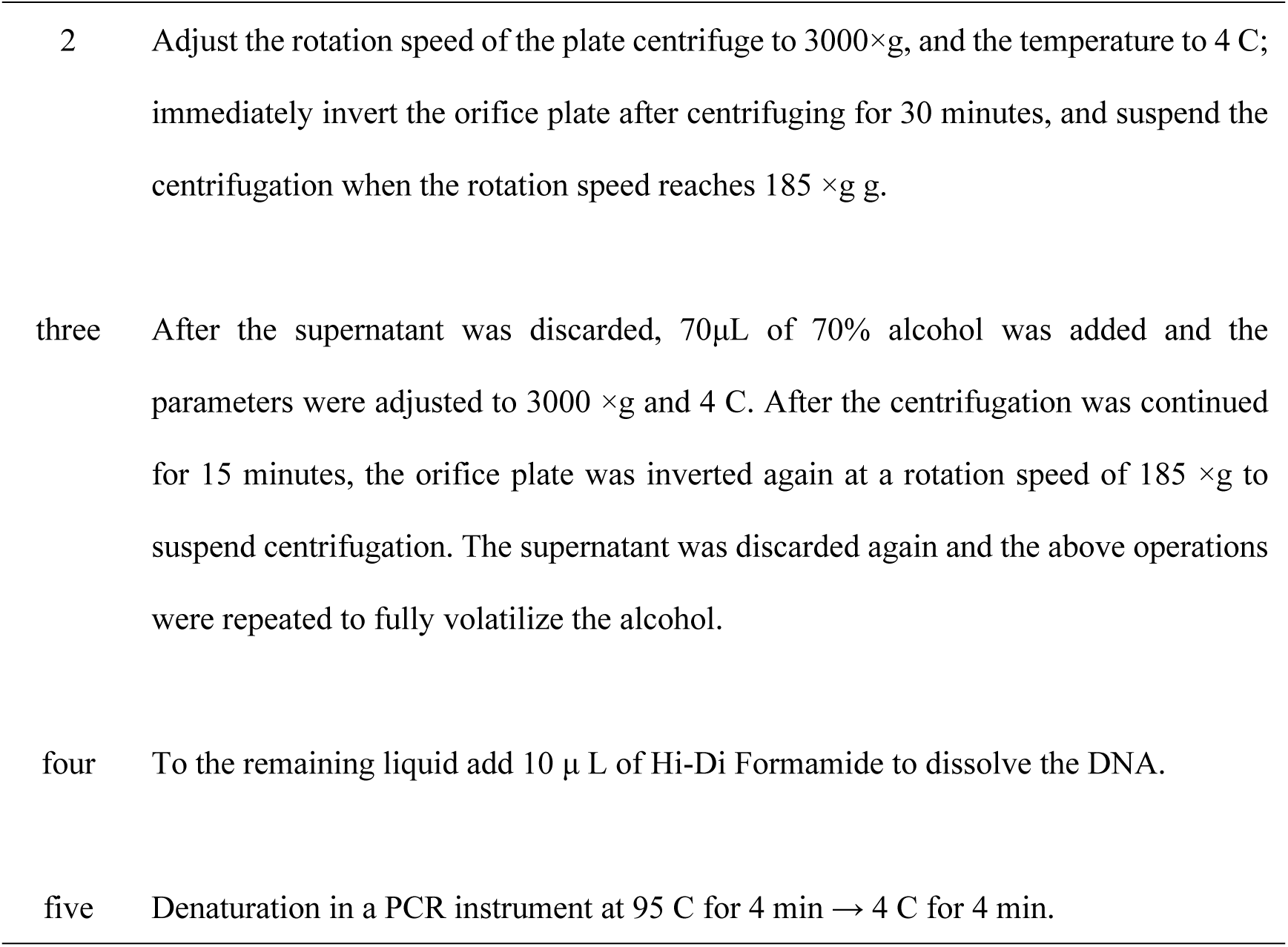
purification of 9: PCR sequence sequencing product.

##### 2.1.11 Sequencing and sequencing results

1. PCR products were sequenced using a 3730XL sequencer produced by ABI Corporation in the United States.
2. The DNA sequencing map generated by sequencing results is generally composed of curves of green (A), blue (C), black (G) and red (T). Heterozygous sites were shown as nested two peaks in the sequencing map, and the resulting sequencing map was read by Chromas software. The genotyping results are shown in the figure below.

**Fig. 6:**
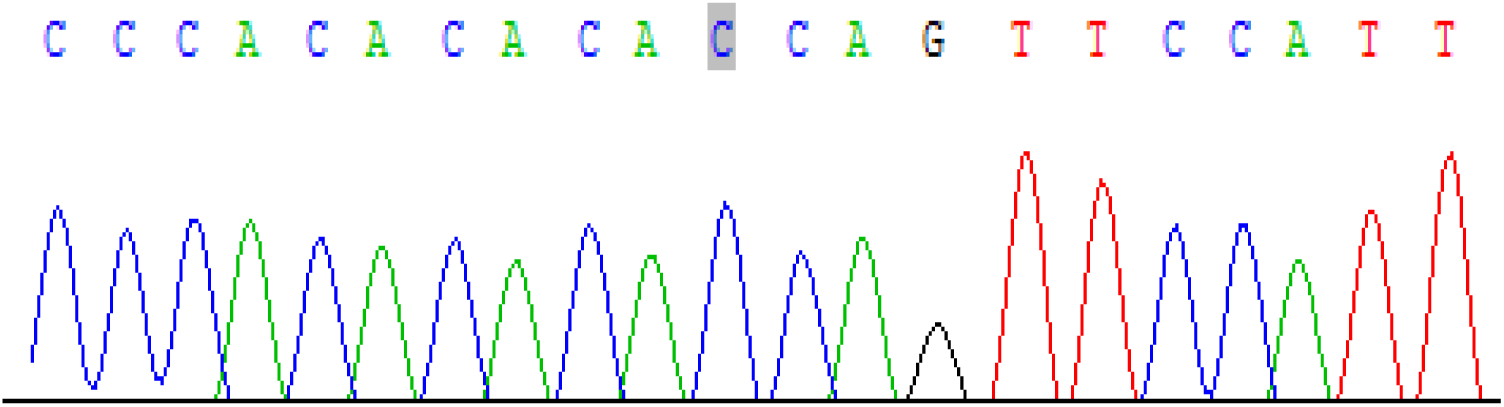
the grey area of the SNP sequencing map of rs473702 of 6: KLF14 gene shows that the genotype at this locus is CC.

**Fig. 7:**
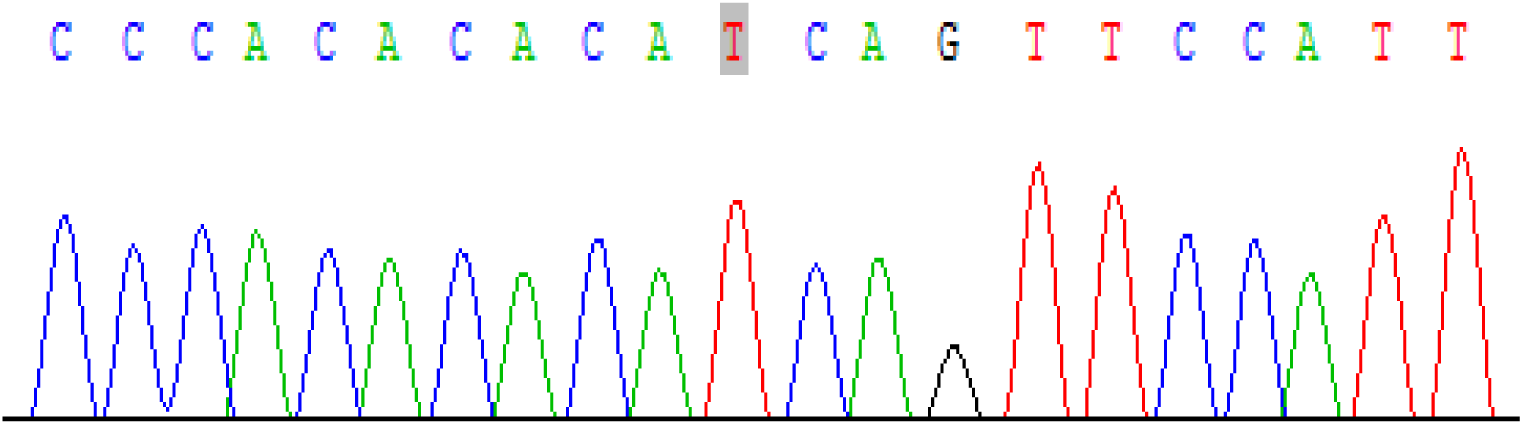
rs4731702SNP sequencing map of 7: KLF14 gene; the grey area indicates that the genotype at this site is TT.

**Fig. 8:**
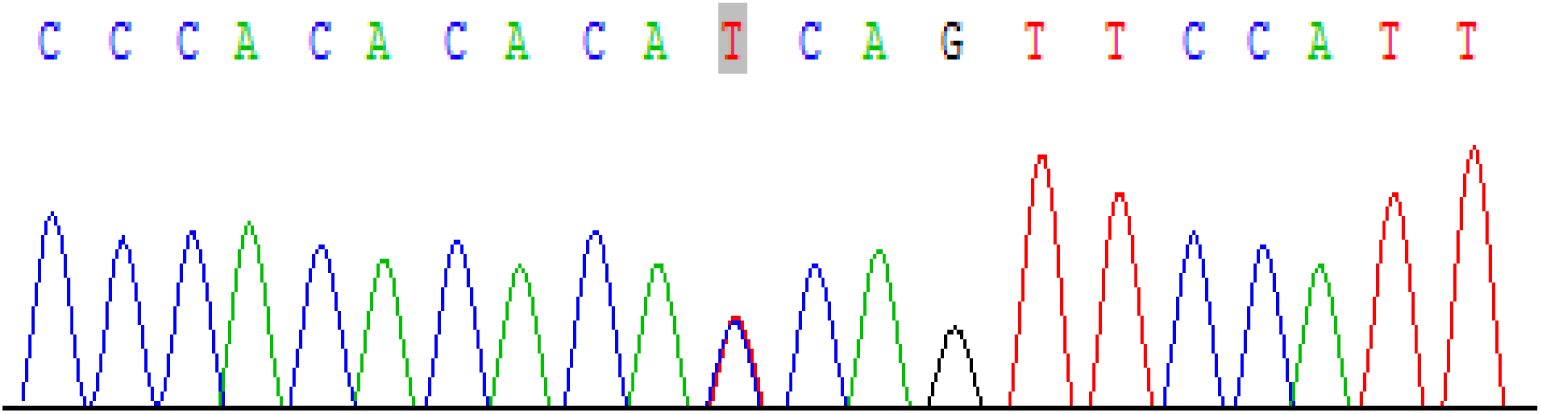
the grey area of the sequencing map of rs4731702SNP of 8: KLF14 gene shows that the genotype at this site is CT.

**Fig. 9:**
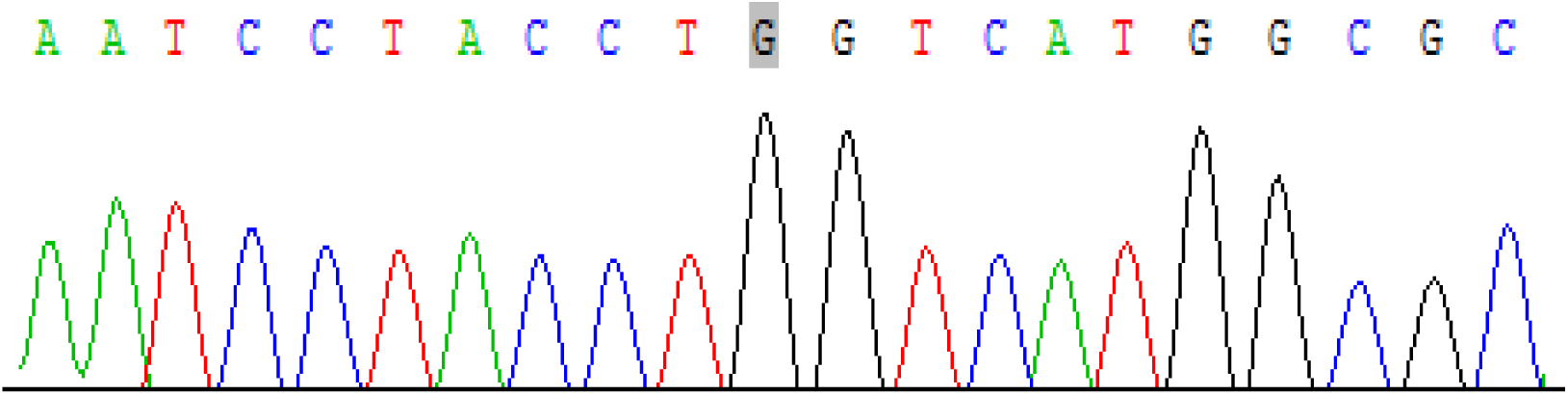
snp sequencing of rs972283 of 9: KLF14 gene the grey area of the map shows that the genotype at this locus is GG.

**Fig. 10:**
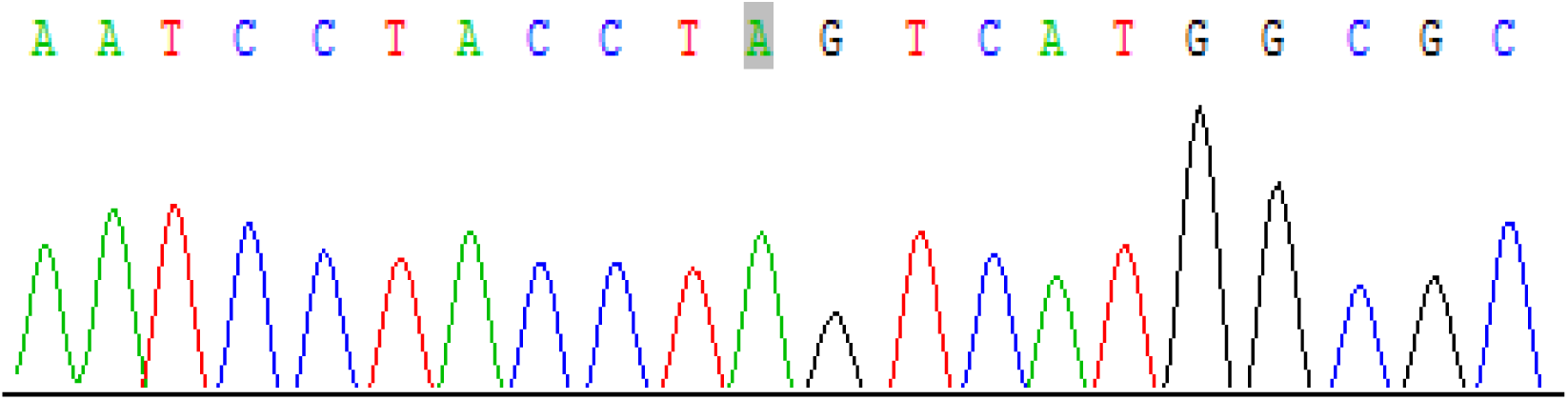
the grey area of the SNP sequencing map of rs972283 of klf14 gene shows that the genotype of this locus is AA.

**Fig. 11:**
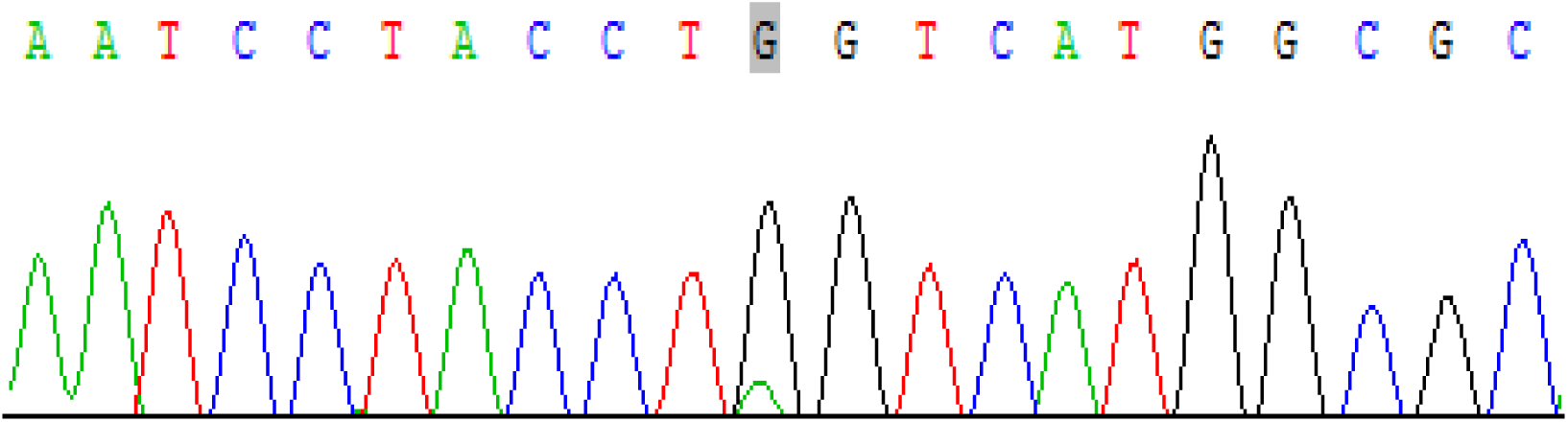
the grey area of the SNP sequencing map of rs972283 of klf14 gene shows that the genotype of this locus is GA.

**Figure 12:**
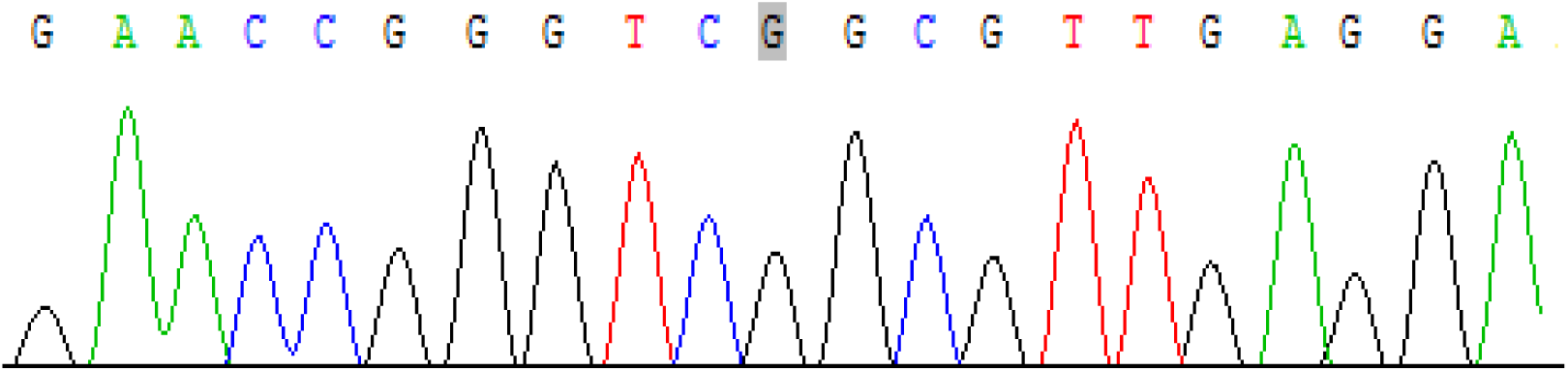
sequence of sr-b1 gene rs5888 that grey area of the sequencing map show that the genotype at this locus was GG.

**Figure 13:**
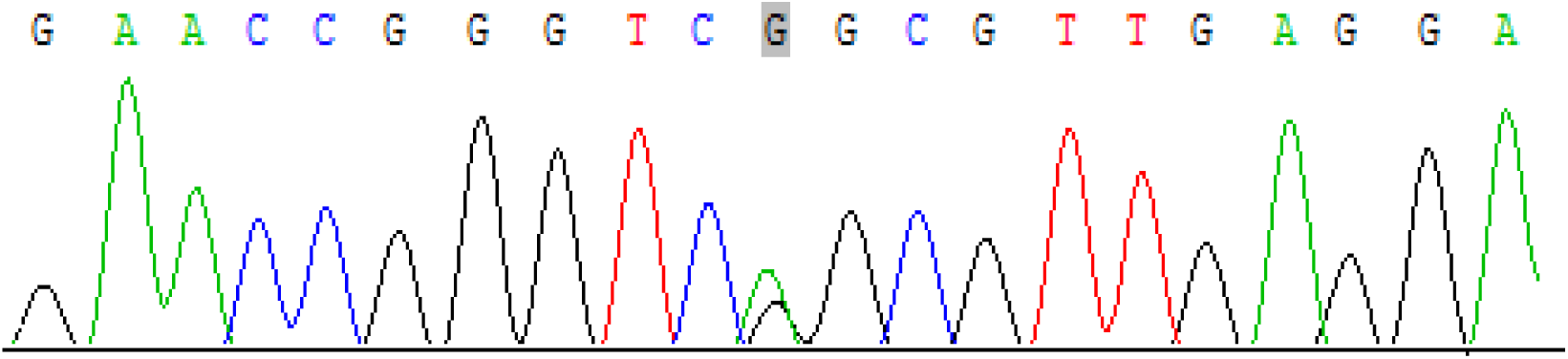
sequence of sr-b1 gene rs5888 that grey area of the sequencing map show that the locus was genotyped as AG.

**Fig. 14:**
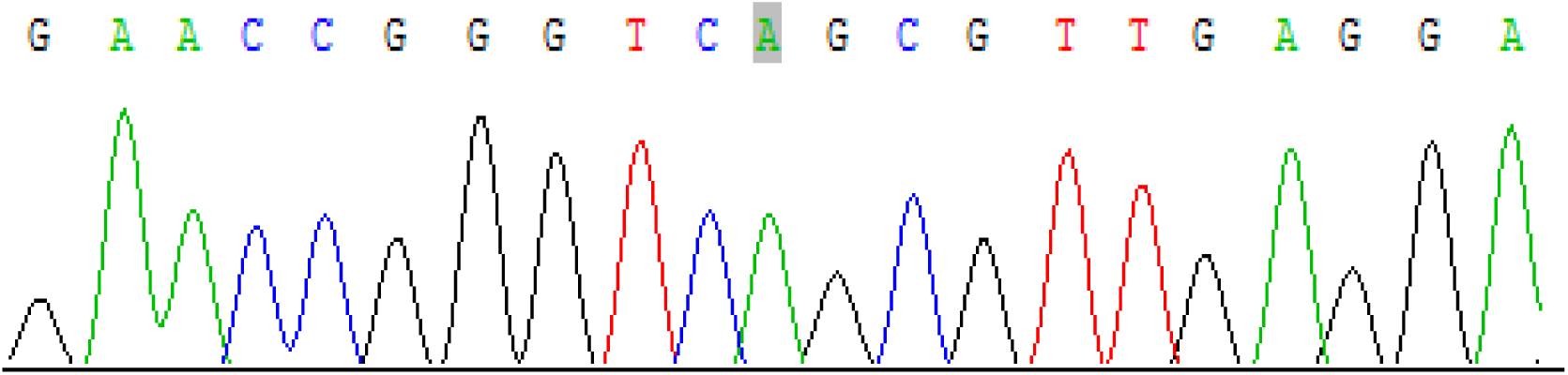
sequence diagram of sr-B1 gene rs5888; grey area indicates that genotype of the locus is AA.

**Fig. 15:**
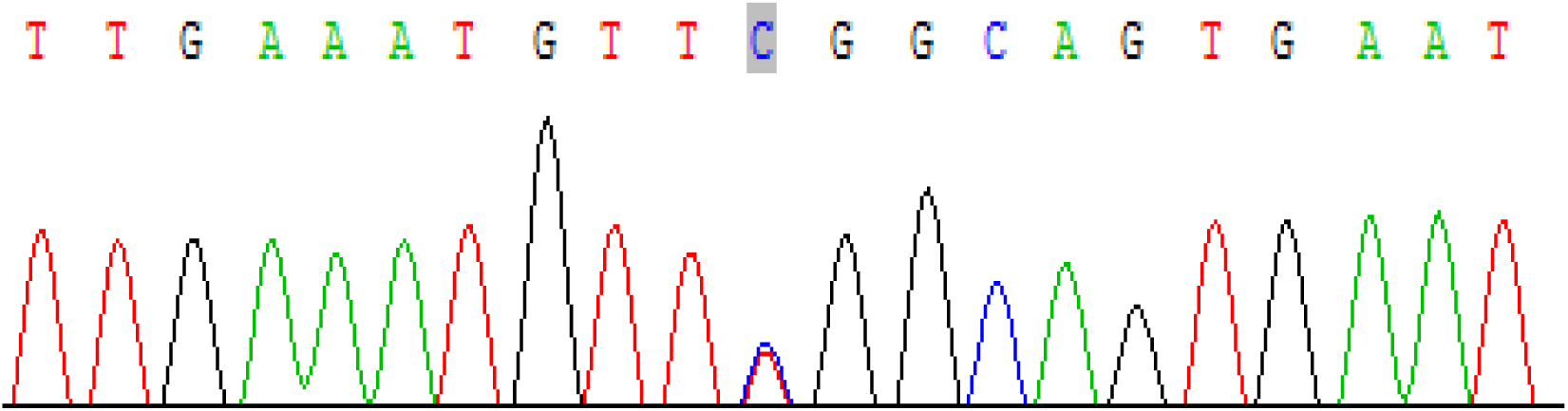
rs838880 sequencing map of SR-B1 gene; the grey area indicates that the genotype at this site is TC.

**Fig. 16:**
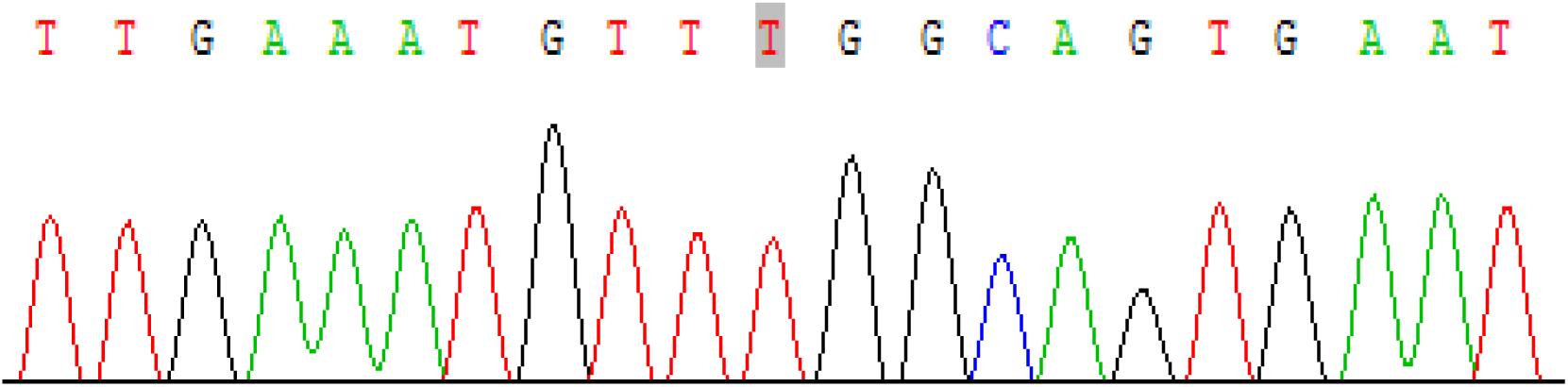
rs838880 sequencing map of SR-B1 gene; the grey area indicates that the genotype at this site is TT.

**Fig. 17:**
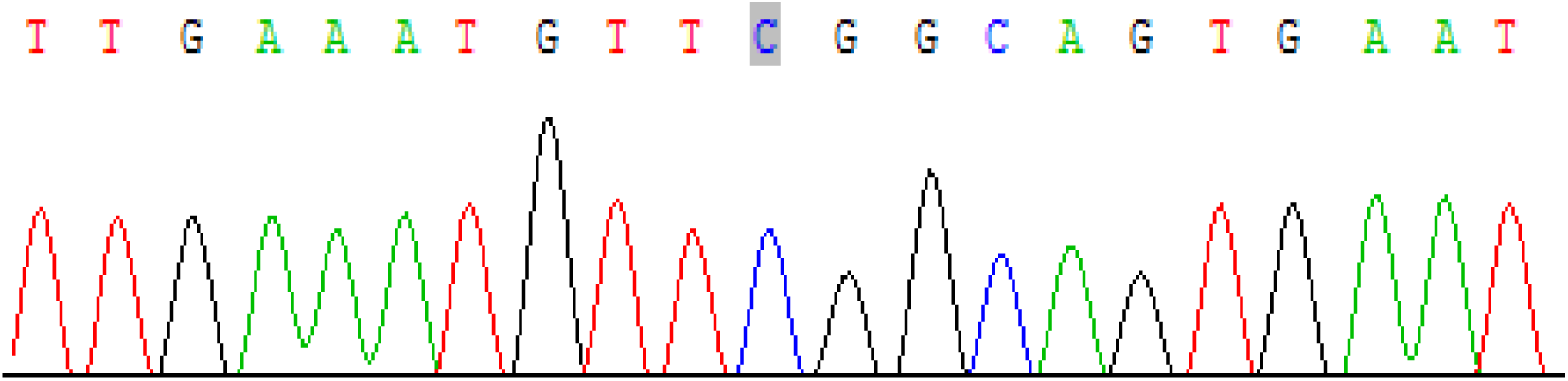
rs838880 sequencing map of SR-B1 gene; the grey area indicates that the genotype at this site is CC.

##### 2.1.12 Chemical qualitative classification of gallstones

Detection method: The gallstones removed during the operation were rinsed several times with distilled water, ground into powder, and a small amount was placed in a 300ml container with 4–5 drops of acetic anhydride and 1–2 drops of concentrated sulfuric acid added, and the color change of the powder was observed. Cholesterol powder was emerald green and radiated from inside to outside. For biliary pigment powder, it is red and emits light from inside to outside.

#### 2.2 Statistical analysis

The experimental data were processed using SPSS 26.0 statistical software. The measurement data in accordance with the normal distribution were expressed as mean standard deviation, and independent sample t test was used. The enumeration data were expressed as percentage (%). The gender differences as well as the genotypic and allelic frequencies of KLF14 (rs4731702 and rs972283) and SR-B1 (rs5888 and rs838880) loci in the case group and the control group were compared with Pearson’s chi-square test. Inter-group comparison was performed using the F test. The risk factors for the development of cholesterol gallstones were analyzed using a binary logistic regression curve, and the two sides of the test level α= 0.05 (P<0.05) indicated that the difference was statistically significant.

## Bear fruit

### 1 Comparison of general data between case group and control group

As shown in Table 8, there was no significant difference between the two groups of general information (P>0.05).

**Table 8:**
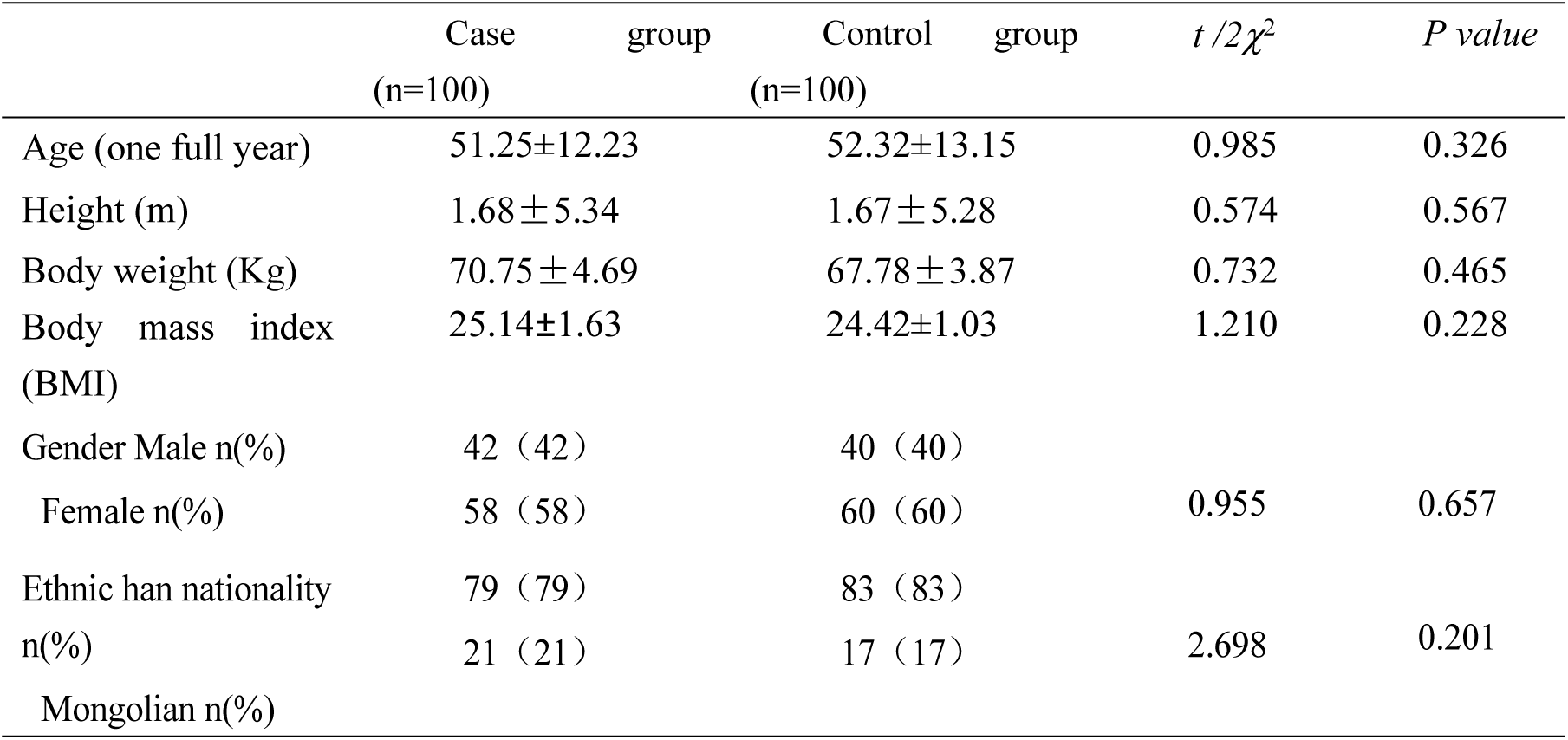
Comparison of General Information between Two Groups.

| | Case group<br>(n=100) | Control group<br>(n=100) | $t/2\chi^2$ | *P value* |
| --- | --- | --- | --- | --- |
| Age (one full year) | 51.25±12.23 | 52.32±13.15 | 0.985 | 0.326 |
| Height (m) | 1.68±5.34 | 1.67±5.28 | 0.574 | 0.567 |
| Body weight (Kg) | 70.75±4.69 | 67.78±3.87 | 0.732 | 0.465 |
| Body mass index<br>(BMI) | 25.14±1.63 | 24.42±1.03 | 1.210 | 0.228 |
| Gender Male n(%) | 42 (42) | 40 (40) | 0.955 | 0.657 |
| Female n(%) | 58 (58) | 60 (60) |  |  |
| Ethnic han nationality<br>n(%) | 79 (79) | 83 (83) | 2.698 | 0.201 |
| Mongolian n(%) | 21 (21) | 17 (17) |  |  |

### 2 Comparison of blood lipid and apolipoprotein between case group and control group

As shown in Table 9, the comparison of blood lipid and lipoprotein indicators between the two groups showed that the Apo-AI/ Apo-B ratio between the case group and the control group was not statistically significant (*P* > 0.05). Compared with the control group, the levels of TG, TCH, LDL and Apo-B in the case group were significantly higher than those in the control group (*P* < 0.05). The HDL and Apo-AI levels in the case group were significantly lower than those in the control group (*P*<0.05), as shown in Figs. 18–19.

**Figure 18:**
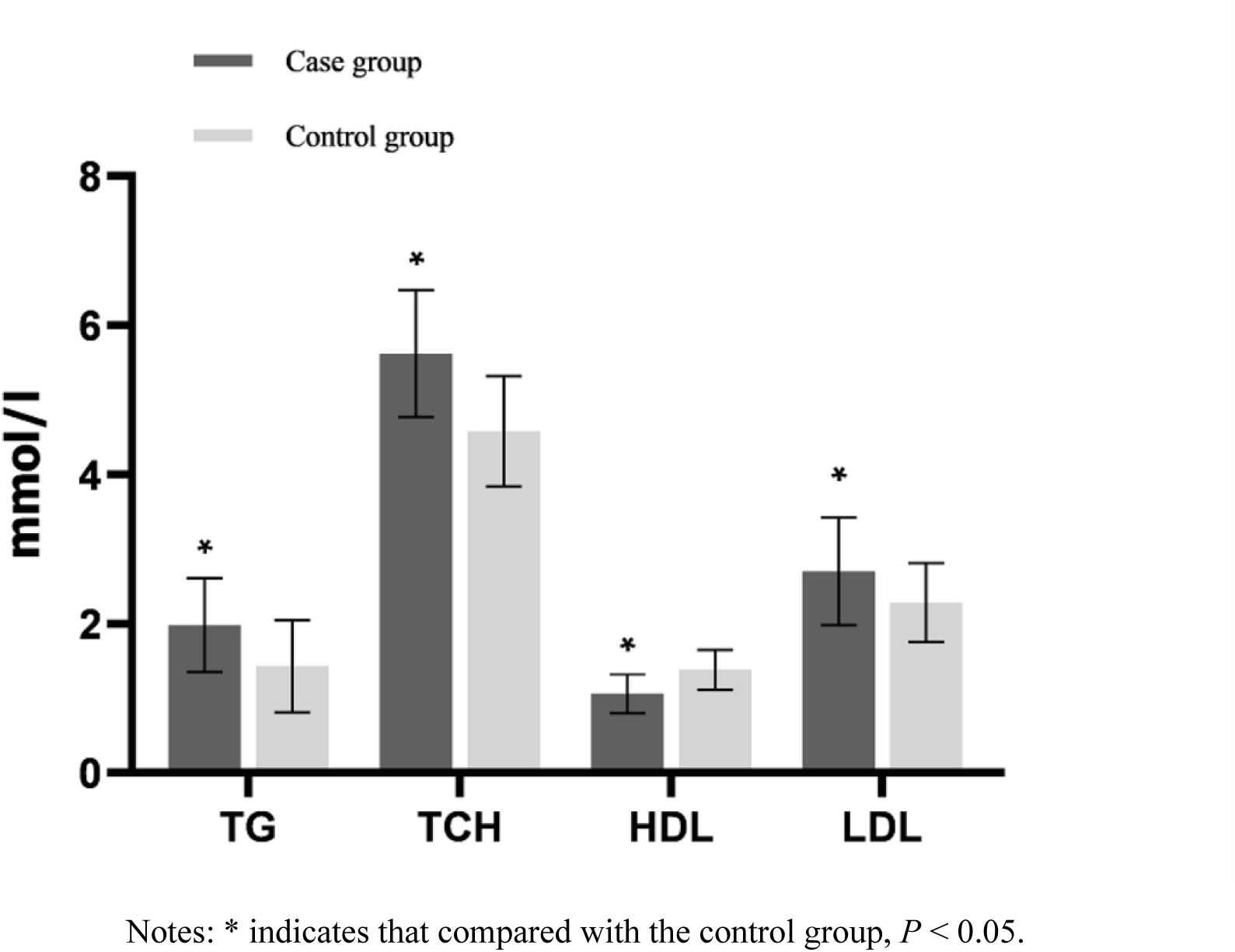
Comparison of blood lipid indexes between two groups.

**Figure 19:**
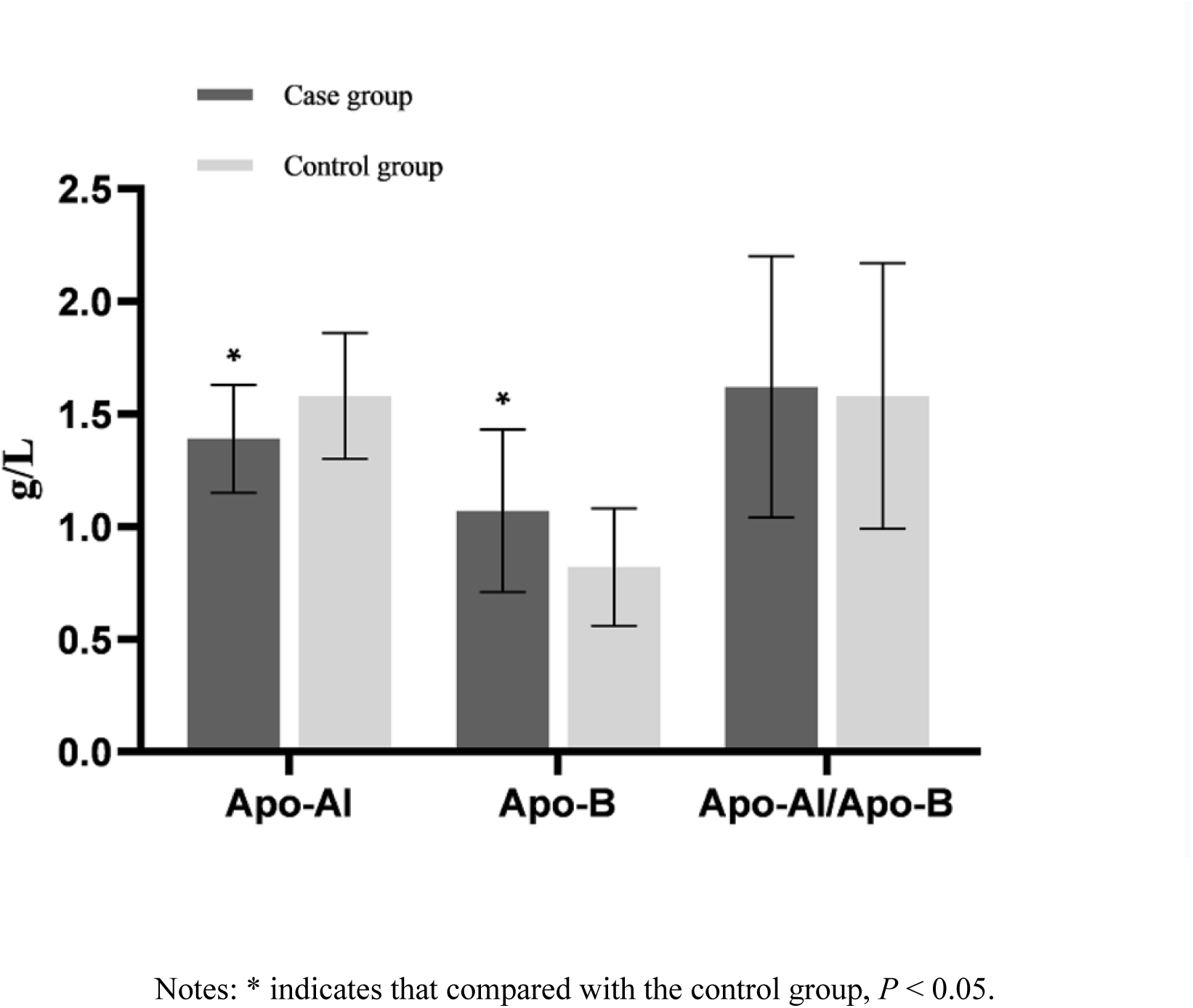
Comparison of blood lipid indexes between two groups.

**Table 9:** Comparison of Blood Lipid Indices between Two Groups.

| Detection Indicator | Case group (100 cases) | Control group (100 cases) | *t* | *P value* |
| --- | --- | --- | --- | --- |
| TG(mmol/L) | 1.98±0.63* | 1.43±0.62 | 2.230 | 0.027 |
| TCH(mmol/L) | 5.62±0.85* | 4.58±0.74 | 2.075 | 0.039 |
| HDL(mmol/L) | 1.06±0.26* | 1.38±0.27 | 2.272 | 0.024 |
| LDL(mmol/L) | 2.70±0.72* | 2.28±0.53 | 2.114 | 0.036 |
| Apo-AI ( g/L ) | 1.39±0.24* | 1.58±0.28 | 2.457 | 0.015 |
| Apo-B ( g/L ) | 1.07±0.36* | 0.82±0.26 | 2.055 | 0.041 |
| Apo-AI/ Apo-B | 1.62±0.58 | 1.58±0.59 | 1.057 | 0.292 |

Notes: \* indicates that compared with the control group, $P < 0.05$ .

## Statistical analysis of rs4731702 at 3 KLF14 gene locus

### 3.1 Comparison of genotypic and allelic frequencies of rs4731702 at the KLF14 locus

The genotypic distribution of the KLF14 gene at rs4731702 in the case group and the control group complied with Hardy-Weinberg’s law of genetic balance (*P* > 0.05), and three genotypes, TT, CT, and CC, could be detected in both groups.

Sample size n≥40, and all T≥5. The Pearson Chi-Square test showed that the gene frequencies of TT, CT AND, and CC in the case group were 10%, 42%, and 48%, respectively, while the genotypic frequencies of 12%, 58%, and 30%, respectively, in the control group. There was a statistically significant difference in the distribution between the two groups (*P* < 0.05). The genotypic frequency of CC in the two groups was higher than that of CT and TT (*P* < 0.05), suggesting that the mutant genotype CC might be a risk factor for cholesterol gallstones (OR = 2.154). The frequencies of alleles C and T in the case group were 69% and 31%, respectively, while those in the control group were 59% and 41%, respectively, showing a statistically significant difference between the two groups (*P* < 0.05). The frequency of allele C was higher than that of allele T, suggesting that allele C might increase the risk of gallbladder cholesterol gallstones (OR = 1.547), as shown in Table 10.

**Table 10:**
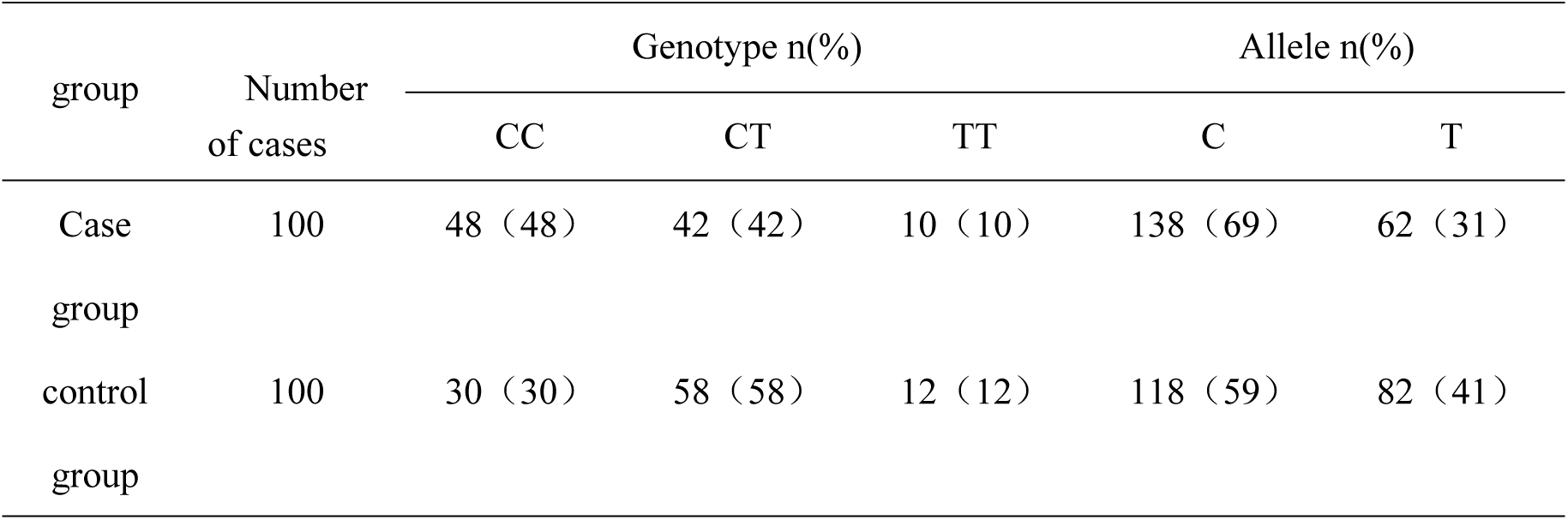
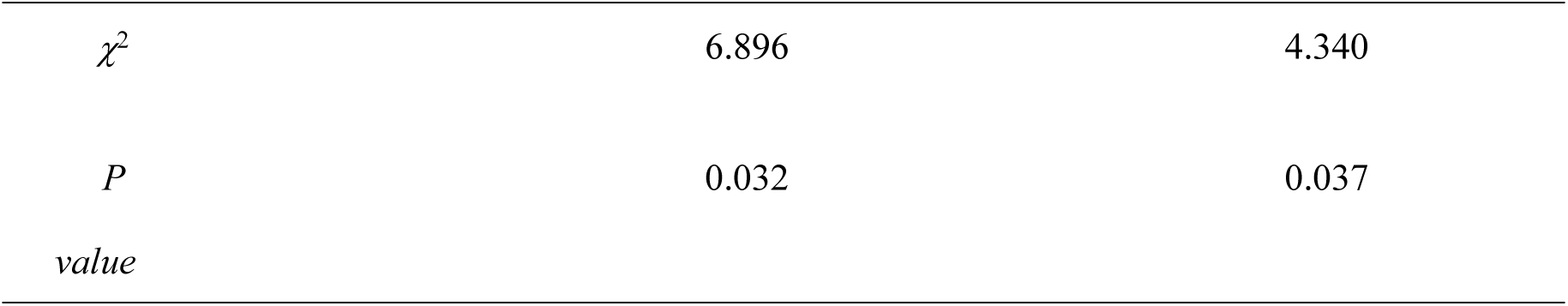
Comparison of Genotypic and Allelic Frequencies of rs4731702 at the 12: KLF14 Locus.

### 3.2 Comparison of clinical data of rs4731702 at KLF14 gene locus in a case group

#### 3.2.1 Comparison of different genotypes of RS473702 with general information

The comparison of genotypes CC, CT and TT within the cholelithiasis group revealed that the ethnic differences were not statistically significant (*P*>0.05), while the gender differences were statistically significant (*P*<0.05), as shown in Table 11.

**Table 11:** Comparison of rs4731702 Genotype at KLF14 Gene Locus with General Information.

| project | genotype | | $\chi^2$ | $P$ value |
| --- | --- | --- | --- | --- |
|  | CC | CT+TT |  |  |
| Gender Male n(%) | 12 (25) | 30 (58) | 10.951 | 0.001 |
| Female n(%) | 36 (75) | 22 (42) |  |  |
| Ethnic han nationality n(%) | 39 (81) | 40 (77) | 0.282 | 0.596 |
| Mongolian n(%) | 9 (19) | 12 (23) |  |  |

#### 3.2.2 Comparison of different genotypes of RS473702 with blood lipid indicators

The TG level of CC genotype in the case group was higher than those of CT and TT genotype, and the difference was statistically significant (*P* < 0.05). The levels of HDL, Apo-AI and Apo-AI/ Apo-B in CC genotype were lower than those in CT and TT genotype, and the differences were statistically significant (*P* < 0.05). There was no significant difference in TCH, LDL or Apo-B (*P* > 0.05), as shown in Table 12.

**Table 12:** Comparison of rs4731702 Genotype and Blood Lipid Indices at KLF14 Gene Locus.

| Detection<br>Indicator | genotype |  |  | *F* | *P value* |
| --- | --- | --- | --- | --- | --- |
|  | CC (n=48) | CT (n=42) | TT (n=10) |  |  |
| TG(mmol/L) | 3.76±1.62* | 2.89±1.25 | 2.60±1.36 | 4.210 | 0.035 |
| TCH(mmol/L) | 7.18±1.63 | 7.06±1.24 | 7.46±1.46 | 1.257 | 0.578 |
| HDL(mmol/L) | 2.08±0.58* | 2.67±0.62 | 2.87±0.27 | 4.984 | 0.017 |
| LDL(mmol/L) | 4.07±0.97 | 3.73±1.29 | 3.66±1.62 | 1.752 | 0.358 |
| Apo-AI (g/L) | 1.21±0.68* | 2.17±0.52 | 2.07±0.37 | 4.746 | 0.022 |
| Apo-B (g/L) | 1.28±0.46 | 1.32±0.65 | 1.22±0.26 | 0.974 | 0.762 |
| Apo-AI/ Apo-B | 2.02±0.51* | 2.89±0.68 | 2.83±0.37 | 4.412 | 0.029 |

Notes: \* Indicates $P < 0.05$ compared with CT and TT genotypes.

## Statistical analysis of rs972283 at 4 KLF14 gene locus

### 4.1 Comparison of genotypic and allelic frequencies of rs972283 at the KLF14 locus

The genotypic distribution at rs972283 of the KLF14 gene in the case group and the control group complied with Hardy-Weinberg’s law of genetic balance (*P* > 0.05), and three genotypes, GG, GA and AA, could be detected in both groups. With the sample size n≥40 and all T≥5, Pearson Chi-Square test was selected and the results showed that the gene frequencies of GG, GA and AA in the case group were 44%, 43% and 13%, respectively, while those of the control group were 45%, 39% and 16%, respectively. There was no significant difference in the distribution between the two groups (*P* > 0.05). The frequencies of allele G and allele A in the case group were 66% and 34%, respectively, while those in the control group were 65% and 35%, respectively. There was no significant difference in the distribution between the two groups (*P* > 0.05), as shown in Table 13.

**Table 13:** Comparison of Genotypic and Allele Frequencies at the 13: KLF14 Locus rs972283.

| group | Number of cases | Genotype n(%) |  |  | Allele n(%) |  |
| --- | --- | --- | --- | --- | --- | --- |
|  |  | GG | GA | AA | G | A |
| Case group | 100 | 44 (44) | 43 (43) | 13 (13) | 131 (66) | 69 (34) |
| control group | 100 | 45 (45) | 39 (39) | 16 (16) | 129 (65) | 71 (35) |
| $\chi^2$ | | | 0.517 | | | 0.044 |
| $P$ | | | 0.772 | | | 0.834 |
| value |  |  |  |  |  |  |

### 4.2 Comparison of clinical data of rs972283 at KLF14 gene locus in case group

#### 4.2.1 Comparison of different genotypes of RS 972283 with general information

The comparison of genotypes GG, GA and AA within the cholelithiasis group revealed that there were no significant differences in ethnicity or gender (*P*>0.05), as shown in Table 14.

**Table 14:**
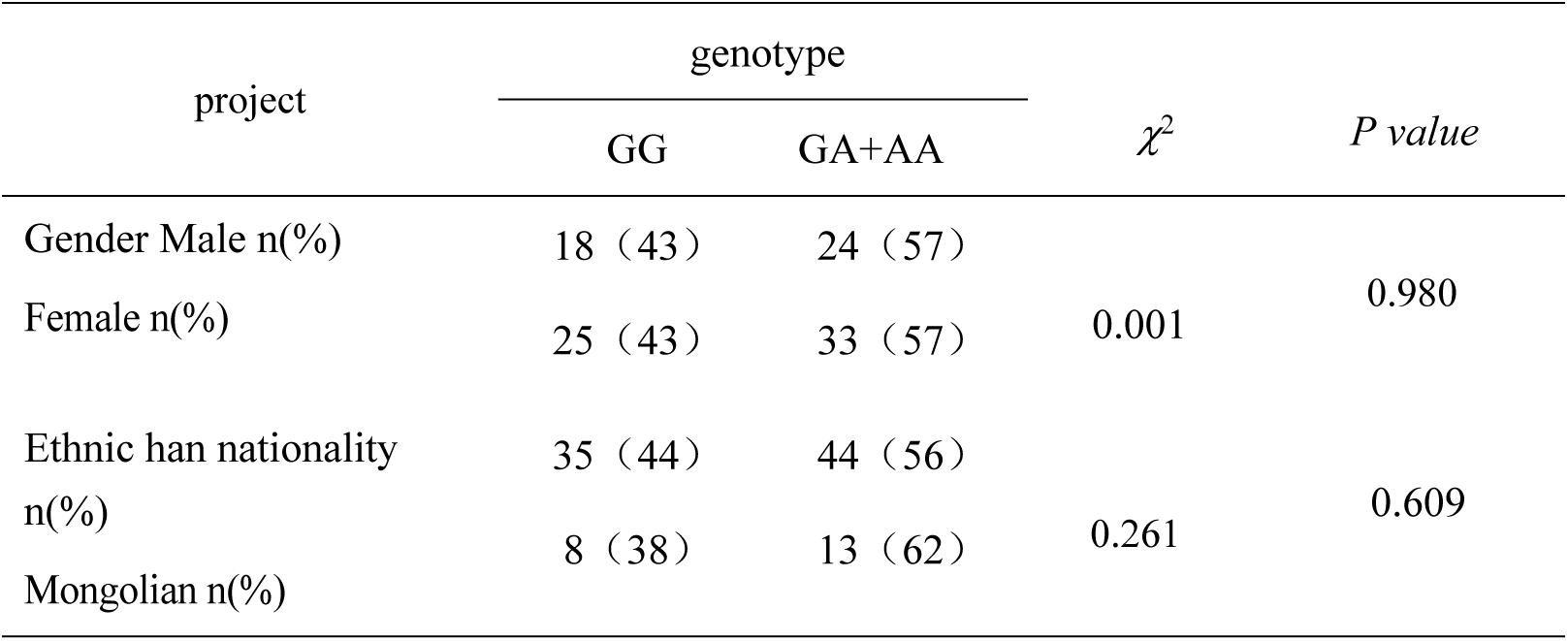
Comparison of rs972283 Genotype at KLF14 Gene Locus with General Information.

| project | genotype | | $\chi^2$ | *P* value |
| --- | --- | --- | --- | --- |
|  | GG | GA+AA |  |  |
| Gender Male n(%) | 18 (43) | 24 (57) | 0.001 | 0.980 |
| Female n(%) | 25 (43) | 33 (57) |  |  |
| Ethnic han nationality n(%) | 35 (44) | 44 (56) | 0.261 | 0.609 |
| Mongolian n(%) | 8 (38) | 13 (62) |  |  |

#### 4.2.2 Comparison of different genotypes of RS972283 with blood lipid indicators

The levels of TG and TCH of GG genotype in the case group were higher than those of GA and AA genotype, and the differences were statistically significant (*P* < 0.05). The levels of HDL, Apo-AI and Apo-AI/ Apo-B in GG genotype were lower than those in GA and AA genotypes, and the differences were statistically significant (*P* < 0.05). There was no significant difference in LDL and Apo-B levels (*P* > 0.05), as shown in Table 15.

**Table 15:** Comparison of rs972283 Genotype and Blood Lipid Indices at KLF14 Locus.

| Detection Indicator | genotype |  |  | *F* | *P* value |
| --- | --- | --- | --- | --- | --- |
|  | GG (n=44) | GA (n=43) | AA (n=13) |  |  |
| TG(mmol/L) | 3.28±1.52* | 2.09±1.35 | 1.96±1.37 | 3.867 | 0.042 |
| TCH(mmol/L) | 6.58±1.73* | 4.76±1.21 | 4.46±1.26 | 4.133 | 0.034 |
| HDL(mmol/L) | 1.88±0.76* | 2.74±0.72 | 2.92±0.37 | 5.210 | 0.015 |
| LDL(mmol/L) | 3.87±0.94 | 3.66±1.39 | 3.69±1.22 | 1.655 | 0.392 |
| Apo-AI (g/L) | 1.07±0.67* | 2.37±0.12 | 2.27±0.27 | 4.849 | 0.021 |
| Apo-B (g/L) | 1.38±0.46 | 1.35±0.61 | 1.29±0.46 | 1.032 | 0.740 |
| Apo-AI/ Apo-B | 1.89±0.41* | 2.79±0.58 | 2.82±0.36 | 4.407 | 0.029 |

Notes: \* Indicates $P < 0.05$ compared with GA and AA genotypes.

## Statistical analysis of rs5888 in 5 SR-B1 gene locus

### 5.1 Comparison of genotypic and allelic frequencies of rs5888 at the SRB1 locus

The distribution of genotypes at rs5888 of the SR-B1 gene in the case group and the control group complied with Hardy-Weinberg’s law of genetic balance (*P* > 0.05), and three genotypes, CC, CT and TT, could be detected in both groups. With the sample size n≥40 and all T≥5, Pearson Chi-Square test was selected and the results showed that the gene frequencies of CC, CT and TT in the case group were 66%, 29% and 5%, respectively, while those of genotype in the control group were 46%, 45% and 9%, respectively. The difference in the distribution between the two groups was statistically significant (*P* < 0.05). The CC genotype at rs5888 of SR-B1 gene in the case group was significantly higher than that of CT+TT genotype (*P* < 0.05), suggesting that the mutant CC genotype might be a risk factor for cholesterol gallstones (OR = 2.279). The frequencies of alleles C and T in the case group were 80% and 20%, respectively, while those in the control group were 68% and 32%, respectively, showing a statistically significant difference between the two groups (*P* < 0.05). The frequency of allele C was higher than that of allele T, suggesting that allele C might increase the risk of gallbladder cholesterol gallstones (OR = 1.898), as shown in Table 16.

**Table 16.** Comparison of Genotypic and Allele Frequencies of rs5888 at the SR-B1 Locus.

| group | Number of cases | Genotype n(%) |  |  | Allele n(%) |  |
| --- | --- | --- | --- | --- | --- | --- |
|  |  | CC | CT | TT | C | T |
| Case group | 100 | 66 (66) | 29 (29) | 5 (5) | 161 (80) | 39 (20) |
| control group | 100 | 46 (46) | 45 (45) | 9 (9) | 137 (68) | 63 (32) |
| $\chi^2$ | | | 8.174 | | | 7.580 |
| *P* value |  |  | 0.017 |  |  | 0.006 |

### 5.2 Comparison of clinical data of rs5888 at SR-B1 gene locus in case groups

#### 5.2.1 Comparison of different genotypes of RS5888 with general information

Through the comparison of genotypes CC, CT and TT in the cholesterol gallstones group, it was found that there were no significant differences in gender and ethnicity (*P*>0.05), as shown in Table 17.

**Table 17.**
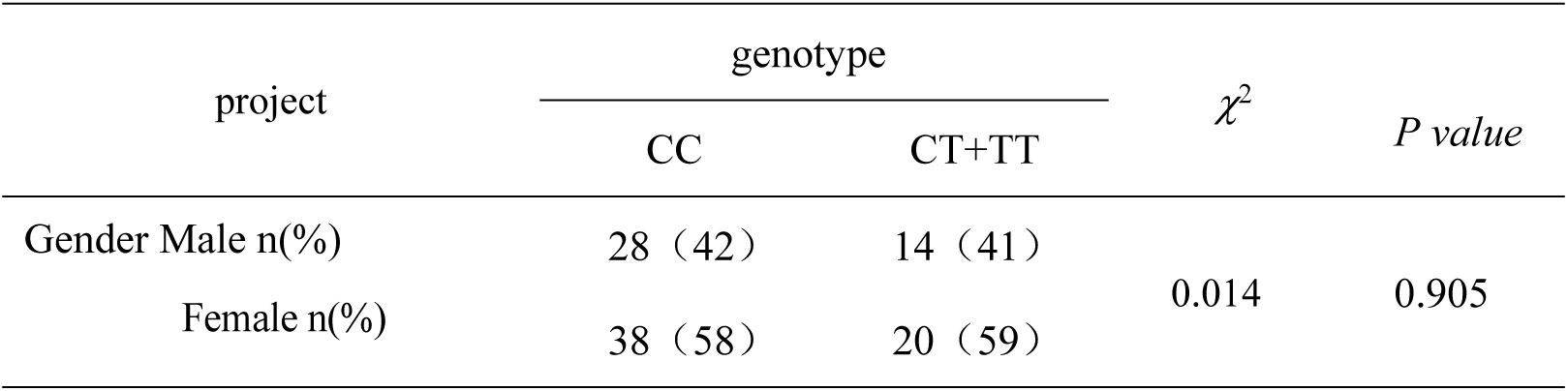
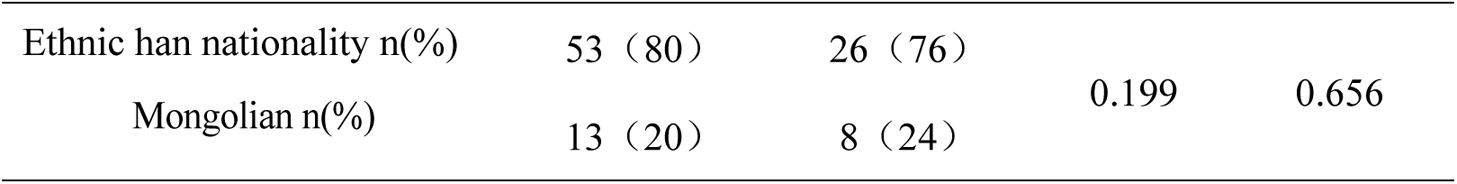
Comparison of rs5888 Genotypes at SR-B1 Locus with General Data.

#### 5.2.2 Comparison of different genotypes of RS5888 and blood lipid indicators

The gene frequency of patients with TT type gene in the case group was significantly lower than that of the control group, and HDL-C of patients with TT type gene was significantly higher than those of CC and CT type, while the contents of TG and TCH were significantly lower than those of CC and CT type (*P* < 0.05). There was no significant difference in LDL, Apo-AI, Apo-B and Apo-AI/ Apo-B (*P* > 0.05), as shown in Table 18.

**Table 18.** Comparison of rs5888 Genotype at SR-B1 Gene Locus and Blood Lipid Indices.

| Lipid Indices |  |  |  |  |  |
| --- | --- | --- | --- | --- | --- |
| Detection Indicator | genotype |  |  | *F* | *P value* |
|  | CC (n=66) | CT (n=29) | TT (n=5) |  |  |
| TG(mmol/L) | 2.11±0.37 | 1.99±0.38 | 1.04±0.38* | 19.953 | 0.000 |
| TCH(mmol/L) | 5.64±0.49 | 5.90±0.38 | 5.69±0.19* | 3.518 | 0.034 |
| HDL(mmol/L) | 1.06±0.14 | 1.05±0.10 | 1.86±0.23* | 82.416 | 0.000 |
| LDL(mmol/L) | 2.72±0.51 | 3.55±0.60 | 3.05±0.19 | 2.237 | 0.112 |
| Apo-AI (g/L) | 1.35±0.19 | 1.37±0.20 | 1.46±0.12 | 0.900 | 0.410 |
| Apo-B (g/L) | 1.07±0.21 | 1.12±0.21 | 1.04±0.26 | 0.469 | 0.627 |
| Apo-AI/ Apo-B | 1.54±0.37 | 1.58±0.38 | 1.79±0.62 | 1.138 | 0.325 |

Notes: \* Indicates $P < 0.05$ compared with CT and TT genotypes.

## Statistical analysis of rs838880 at 6 SR-B1 gene locus

### 6.1 Comparison of genotypic and allelic frequencies of rs838880 at 6.1 SR-B1 locus

The genotype distribution at rs838880 of the SR-B1 gene in the case group and the control group conformed to Hardy-Weinberg’s law of genetic balance (*P* > 0.05), and three genotypes (TC, TT and CC) could be detected in both groups. With the sample size n≥40 and all T≥5, Pearson Chi-Square test was selected and the results showed that the gene frequencies of TC, TT and CC in the case group were 48%, 49% and 3%, respectively, while the genotypic frequencies in the control group were 33%, 36% and 31%, respectively (*P* < 0.05). The number of CC genotype at rs838880 of SR-B1 gene in case group was significantly lower than that in control group, suggesting that genotype CC at rs838880 of SR-B1 gene might be the protective genotype for cholesterol gallstones (OR=0.116). The frequencies of alleles C and T in the case group were 27% and 73%, respectively, while those in the control group were 47% and 53%, respectively. There was a statistically significant difference in the distribution between the two groups (P < 0.05). The frequency of the T allele was higher than that of the C allele, suggesting that the C allele might reduce the risk of cholesterol gallstones (OR = 0.409), as shown in Table 19.

**Table 19:**
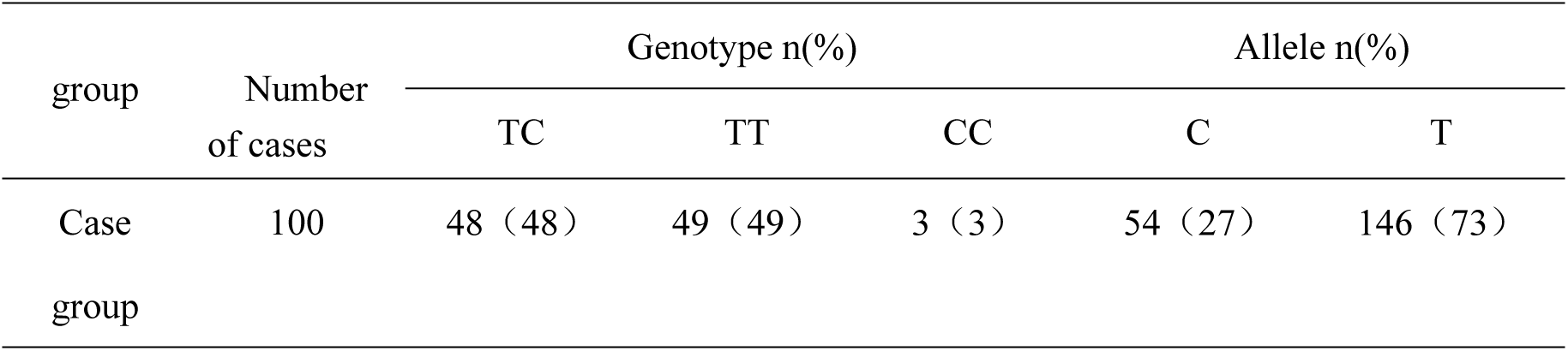
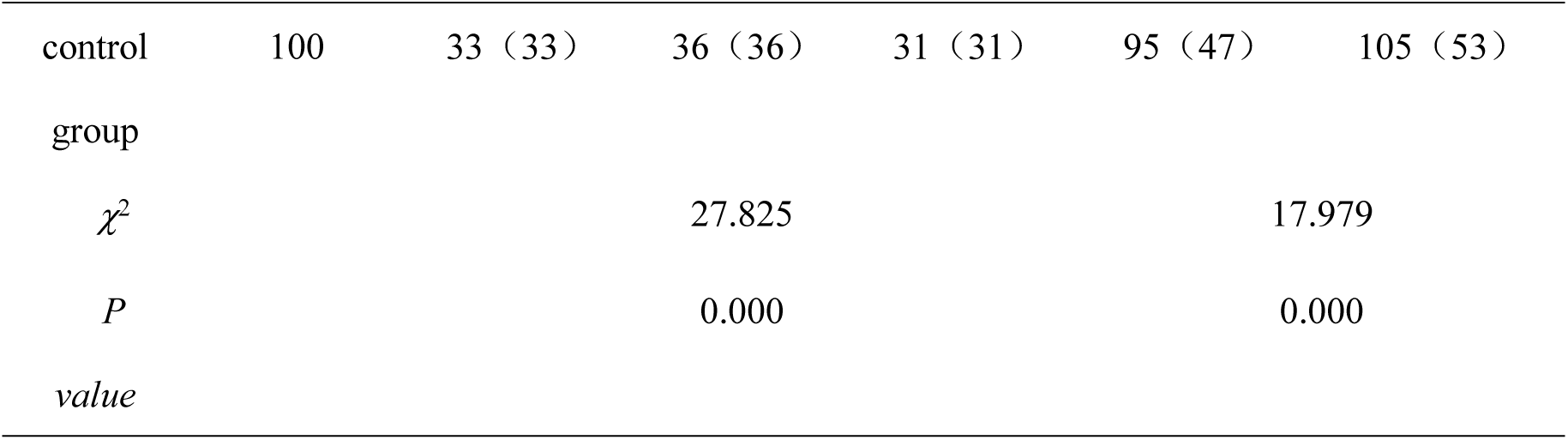
Comparison of Genotypic and Allele Frequencies of rs838880 at the SR-B1 Locus.

### 6.2 Comparison of clinical data of rs838880 at SR-B1 gene locus in case groups

#### 6.2.1 Comparison of different genotypes of RS838880 with general information

The comparison of genotypes TT, TC and CC within the cholelithiasis group revealed that there were no significant differences in ethnicity or gender (*P*>0.05), as shown in Table 20.

**Table 20.** Comparison of rs838880 Genotype at SR-B1 Gene Locus with General Data.

| project | genotype | | $\chi^2$ | *P value* |
| --- | --- | --- | --- | --- |
|  | TT | TC+CC |  |  |
| Gender Male n(%) | 21 (43) | 21 (41) | 0.029 | 0.865 |
| Female n(%) | 28 (57) | 30 (59) |  |  |
| Ethnic han nationality n(%) | 40 (82) | 39 (76) | 0.401 | 0.526 |
| Mongolian n(%) | 9 (18) | 12 (24) |  |  |

#### 6.2.2 Comparison of different genotypes of RS838880 with blood lipid indicators

The gene frequency of CC genotype in the case group was significantly lower than that in the control group. The TCH of patients with TT genotype in the case group was significantly higher than those of patients with TC and CC genotype, and the difference was statistically significant (*P* < 0.05). HDL of patients with CC type gene was significantly higher than those of patients with TC and TT type in this group (*P* < 0.05). This indicates that the C allele may be associated with an increase in HDL-C in blood. There was no significant difference in TG, LDL, Apo-AI and Apo-B (*P* > 0.05), as shown in Table 21.

**Table 21.** Comparison of rs838880 Genotype and Blood Lipid Indices at SR-B1 Gene Locus.

| Detection Indicator | genotype |  |  | *F* | *P value* |
| --- | --- | --- | --- | --- | --- |
|  | TC (n=48) | TT (n=49) | CC (n=3) |  |  |
| TG(mmol/L) | 2.06±0.38 | 1.94±0.41 | 2.25±0.28 | 1.675 | 0.193 |
| TCH(mmol/L) | 5.16±0.27 | 5.96±0.39[1] | 4.95±0.16 | 74.007 | 0.000 |
| HDL(mmol/L) | 1.06±0.17 | 1.05±0.17 | 1.60±0.25[2] | 14.961 | 0.000 |
| LDL(mmol/L) | 2.74±0.43 | 2.74±0.42 | 2.71±0.36 | 0.009 | 0.991 |
| Apo-AI (g/L) | 1.28±0.17 | 1.30±0.21 | 1.44±0.19 | 1.043 | 0.356 |
| Apo-B (g/L) | 1.10±0.23 | 1.09±0.26 | 1.21±0.22 | 0.337 | 0.714 |
| Apo-AI/ Apo-B | 1.85±0.19 | 1.99±0.16[3] | 1.83±0.14 | 8.379 | 0.000 |

Note: [1] indicated that compared with TC and CC genotypes, $P < 0.05$ .
[2] indicated that compared with TC and TT genotypes, $P < 0.05$ .
[3] indicated that compared with TC genotype, $P < 0.05$ .

## Correlation analysis between 7 KLF14 gene and SR-B1 gene

### 7.1 Analysis of association between locus rs4731702 and locus rs972283 of KLF14 gene

Through the association analysis between locus rs4731702 and locus rs972283 of KLF14 gene in case group, we obtained that the column association number was 0.585, and *P*=0.000. It can be considered that although locus rs972283 of KLF14 gene might not be associated with the occurrence of cholesterol gallstones, the above two gene loci may have a certain correlation with the occurrence of cholesterol gallstones, as shown in Table 22.

**Table 22:** cross-table between locus rs4731702 and locus rs972283 of klf14 gene.

| Gene locus and genotype |  | Gene rs972283 |  | amount to |
| --- | --- | --- | --- | --- |
|  |  | GG | GA+AA |  |
| Gene rs4731702 | CC | 39 | nine | 48 |
|  | CT+TT | five | 47 | fifty-two |
| amount to |  | forty-four | fifty-six | 100 |

### 7.2 Correlation analysis between locus rs5888 and locus rs838880 of SR-B1 **gene**

Through the above independent research on the above two genes and analysis on the correlation between rs5888 and rs838880 of SR-B1 gene locus in case group, we have obtained the column contact number is 0.002, and P=0.980. It can be considered that the above two gene loci may play their respective roles in the pathogenesis of gallbladder cholesterol gallstones, but they do not have a certain correlation, as shown in Table 23.

**Table 23:** Cross Table of rs5888 and rs838880 of SR-B1 Gene.

| Gene locus and genotype |  | Gene rs838880 |  | amount to |
| --- | --- | --- | --- | --- |
|  |  | CC | TC+TT |  |
| Gene rs5888 | CC | 2 | 64 | 66 |
|  | CT+TT | one | 33 | 34 |
| amount to |  | three | 97 | 100 |

### 7.3 Correlation analysis between rs5888 of SR-B1 gene locus and rs4731702 of KLF14 gene locus

Through the above studies on the independence of the above two genes respectively and the analysis on the correlation between the rs5888 gene locus of SR-B1 and the rs4731702 gene locus of KLF14 in case group, we have obtained that the column connection number is 0.490, and P=0.000. It can be considered that the above two gene loci may not only play their respective roles in the pathogenesis of gallbladder cholesterol gallstones, but also have a certain correlation, as shown in Table 24.

**Table 24:** Crosstab between SR-B1 locus rs5888 and KLF14 locus rs4731702.

| Gene locus and genotype |  | Gene rs4731702 |  | amount to |
| --- | --- | --- | --- | --- |
|  |  | CC | CT+TT |  |
| Gene rs5888 | CC | 45 | 21 | 66 |
|  | CT+TT | three | 31 | 34 |
| amount to |  | 48 | fifty-two | 100 |

### 7.4 Correlation analysis between rs838880 of SR-B1 gene locus and rs972283 of KLF14 gene locus

Through the correlation analysis of SR-B1 gene locus rs838880 and KLF14 gene locus rs972283 in case group, we obtained the column contact number was 0.038, and *P*=0.706, suggesting that the above two gene loci had no correlation in the pathogenesis of cholesterol gallstones, as shown in Table 25.

**Table 25:** Crosstab between SR-B1 locus rs838880 and KLF14 locus rs972283.

| Gene locus and genotype |  | Gene rs972283 |  |  |
| --- | --- | --- | --- | --- |
|  |  | GG | GA+AA | amount to |
| Gene rs838880 | CC | one | 2 | three |
|  | TC+TT | 43 | 54 | 97 |
| amount to |  | forty-four | fifty-six | 100 |

## 8 multivariate conditional logistic regression analysis

Through the application of multivariate logistic regression model fitting, common environmental risk factors were analyzed and conclusions were drawn, as shown in Table 26. BMI(OR=2.758) and family history of gallbladder cholesterol (OR=85.796) were the risk factors for gallbladder cholesterol (*P* < 0.05). Exercise habit (OR=0.025) was the protective factor for cholesterol gallstones (*P* < 0.05), as shown in Fig. 20.

**Figure 20:**
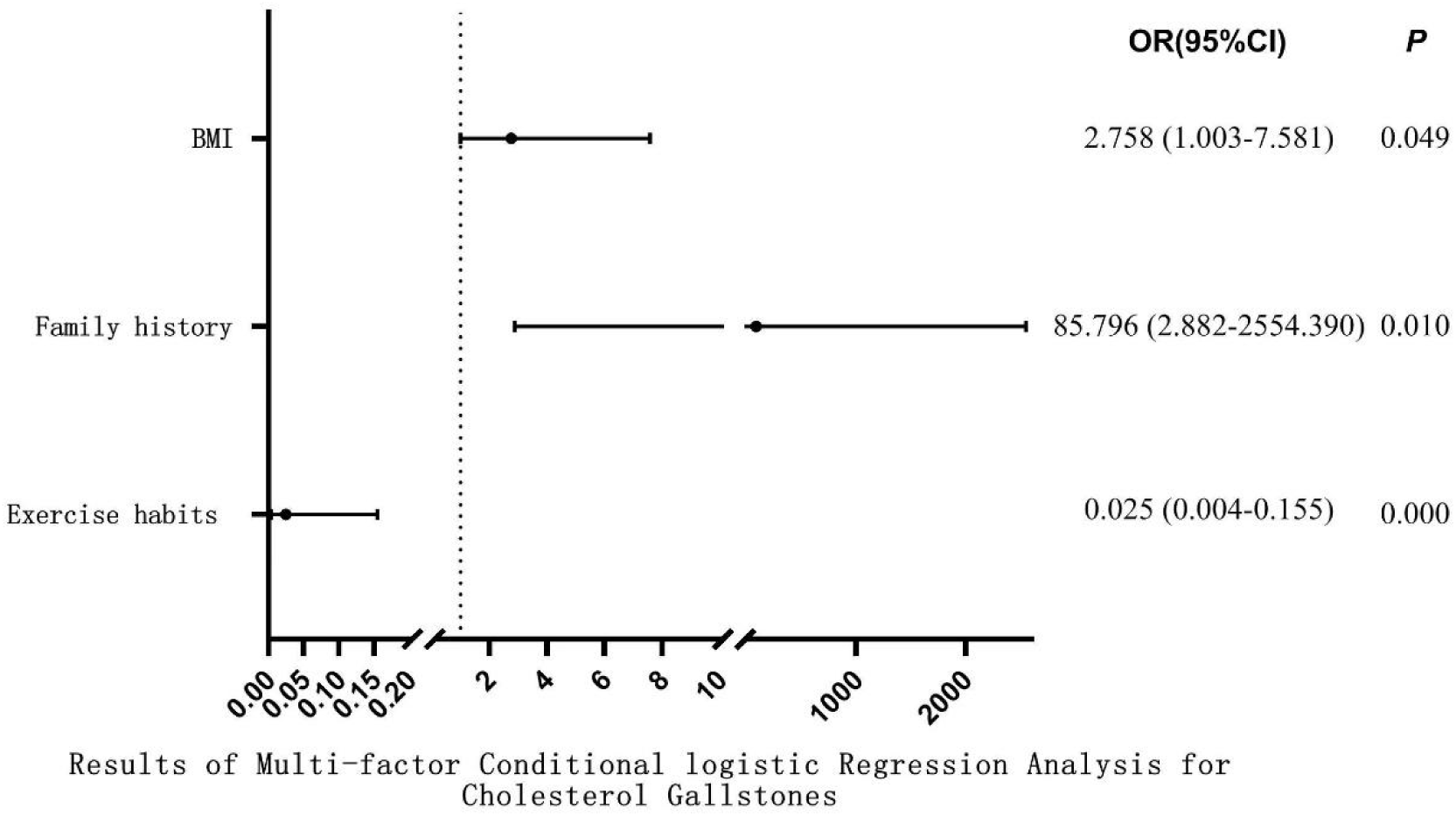
Results of Multi-factor Conditional logistic Regression Analysis for Cholesterol Gallstones.

**Table 26:**
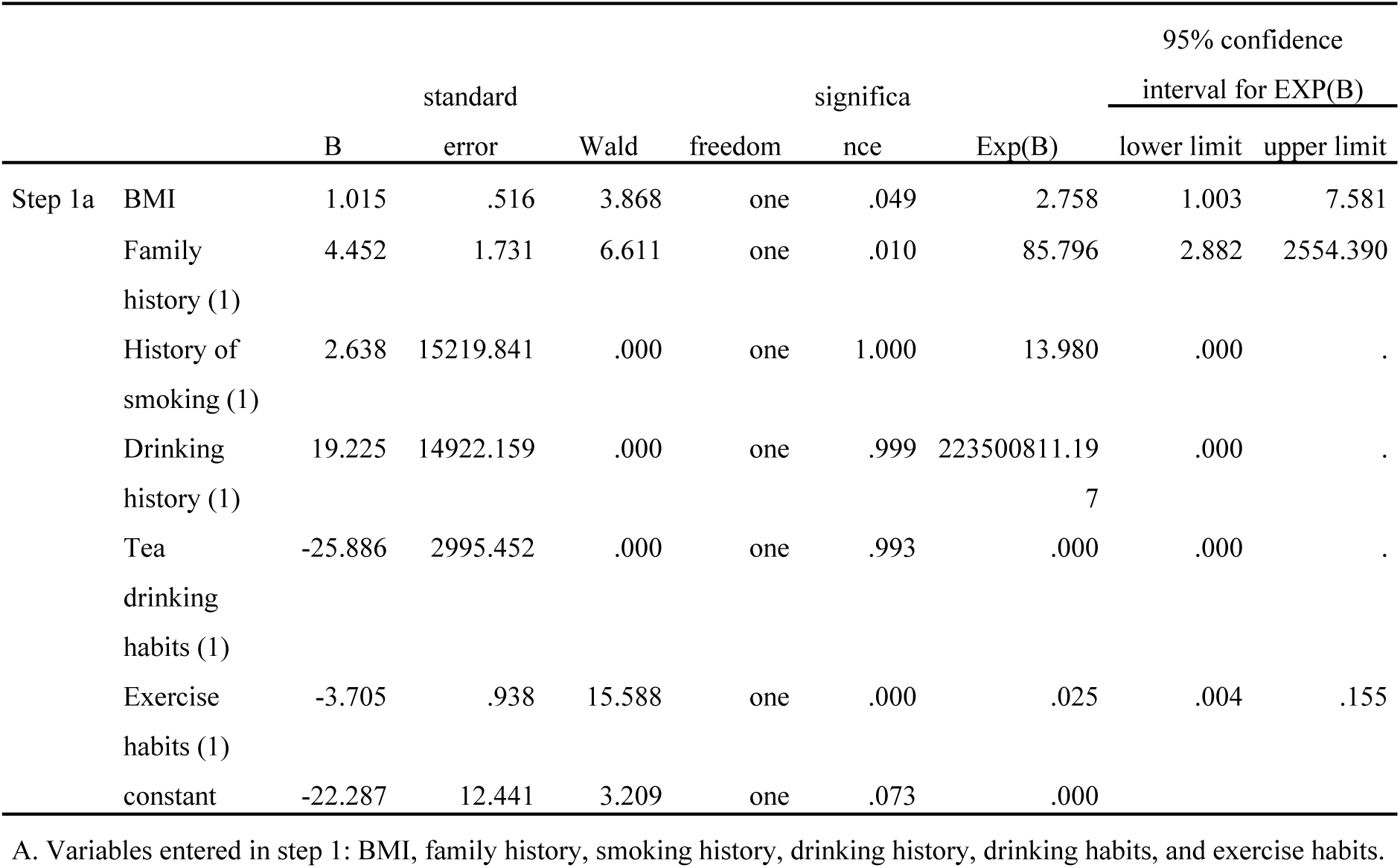
Results of Multi-factor Conditional logistic Regression Analysis for Cholesterol Gallstones.

## 9 gene and environmental factors correlation analysis

No definite conclusion has been drawn in the etiological study of cholesterol gallstones, which is currently generally considered to be the result of the combined action of genes and multiple environmental factors. Through case arrangement and analysis, three environmental factors including BMI, family history of gallstones and exercise habit were selected in the present study to analyze and discuss the association between these three environmental factors and genes. The results are shown in Table 27(*P* < 0.05).

**Table 27:**
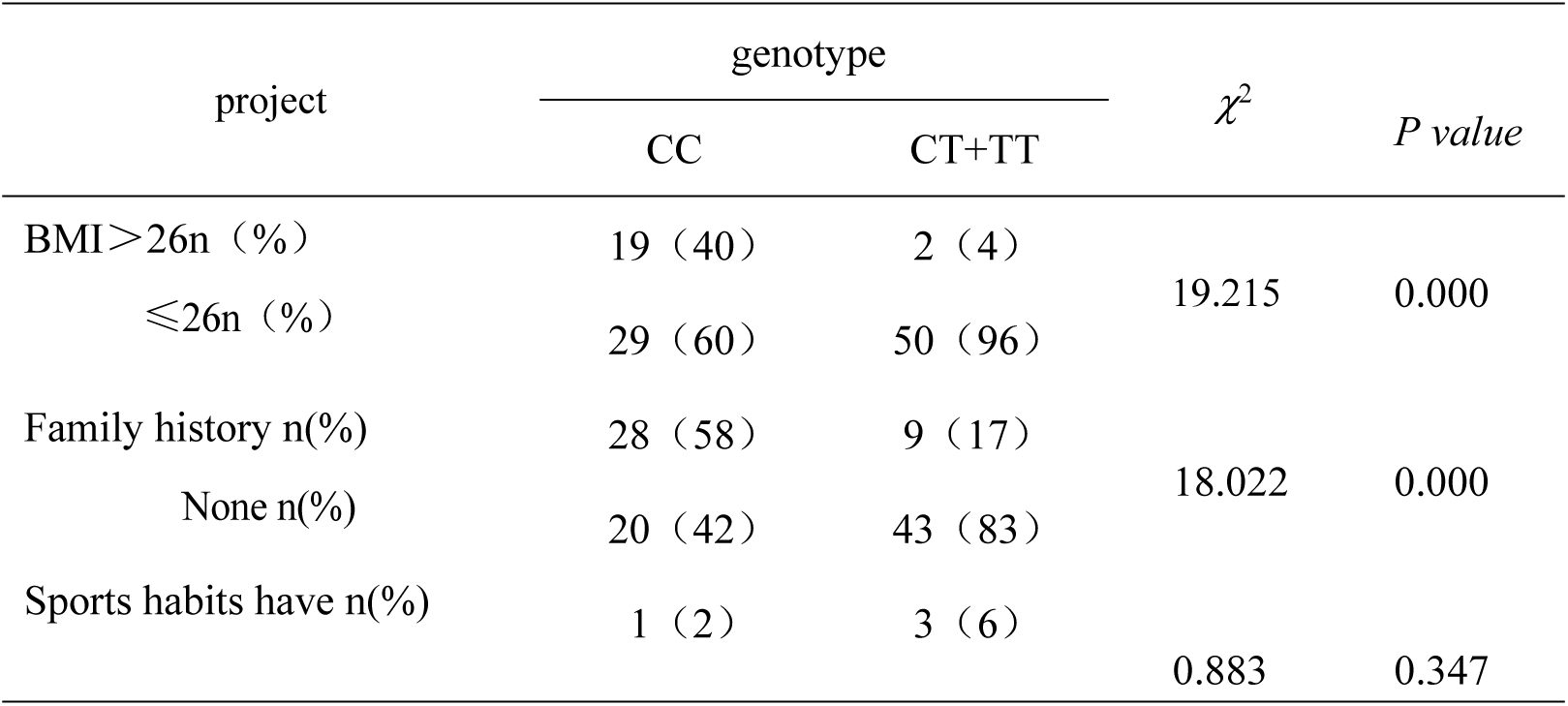
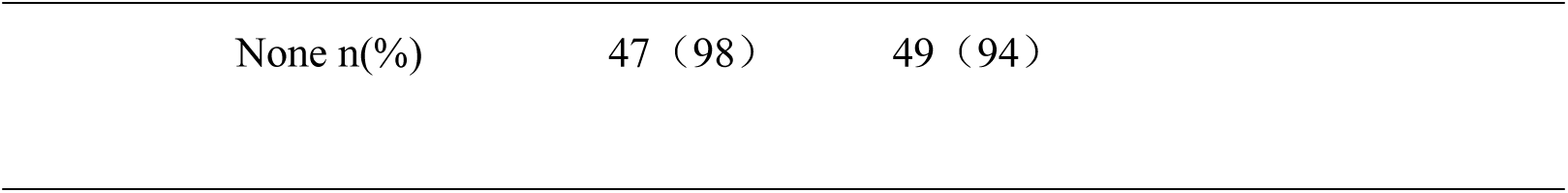
genotype and environmental factor analysis of rs4731702 at klf14 locus.

| project | genotype | | $\chi^2$ | P value |
| --- | --- | --- | --- | --- |
|  | CC | CT+TT |  |  |
| BMI>26n (%) | 19 (40) | 2 (4) | 19.215 | 0.000 |
| ≤26n (%) | 29 (60) | 50 (96) |  |  |
| Family history n(%) | 28 (58) | 9 (17) | 18.022 | 0.000 |
| None n(%) | 20 (42) | 43 (83) |  |  |
| Sports habits have n(%) | 1 (2) | 3 (6) | 0.883 | 0.347 |

|  |  |  |
| --- | --- | --- |
| None n(%) | 47 (98) | 49 (94) |

Through the correlation analysis between RS473702 of KLF14 gene locus and BMI, family history of gallbladder cholesterol, and exercise habit, it was concluded that patients with CC genotype (KLF14 gene RS473702) with BMI > 26 were more likely to suffer from gallbladder cholesterol gallstones (OR=16.379) than BMI < 26 (Table 28). Patients with a family history of gallstones with genotype (KLF14 rs4731702)CC were 6.689-fold (OR=6.689) more likely to have CC than patients without a family history of gallstones with genotype (KLF14 RS 473702) (all *P* < 0.05), as shown in Table 29. There was no statistical significance (*P* > 0.05) of exercise habits or not in patients with genotype (KLF14 gene rs4731702)CC (Table 27).

**Table 28:**
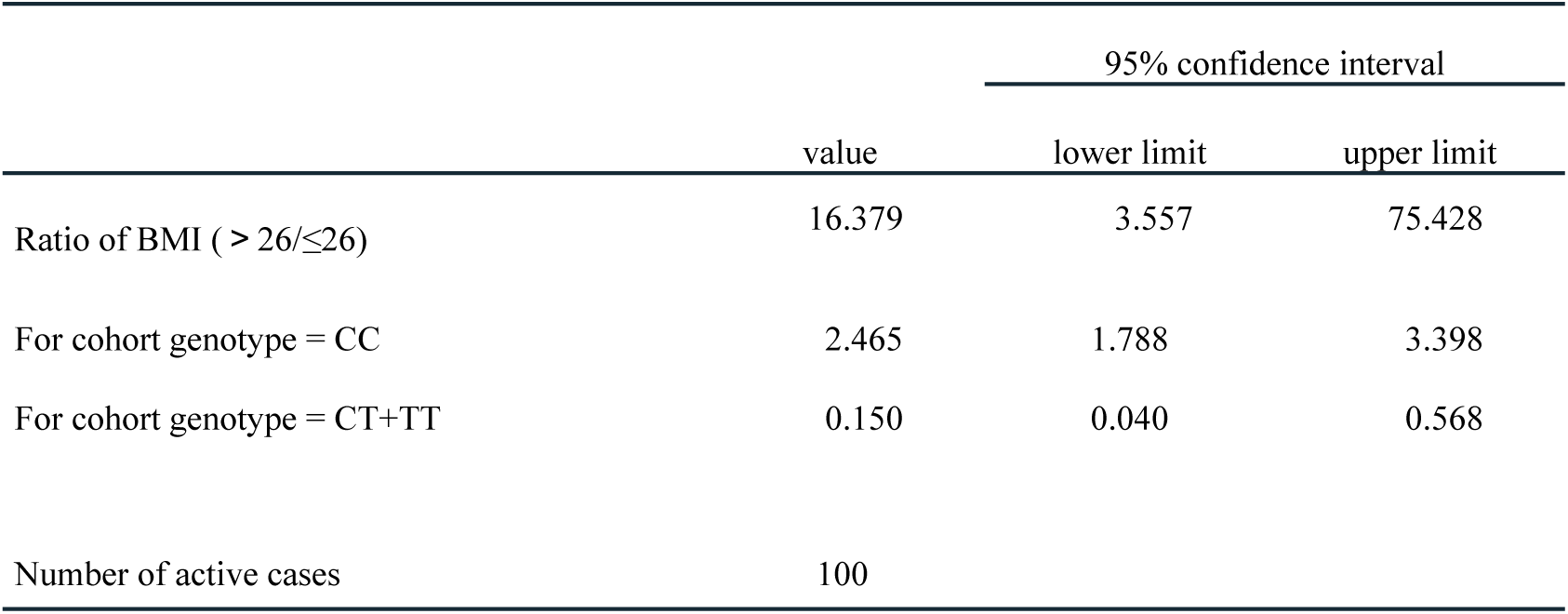
bmi risk assessment.

|  | value | 95% confidence interval |  |
| --- | --- | --- | --- |
|  |  | lower limit | upper limit |
| Ratio of BMI ($> 26/\leq 26$) | 16.379 | 3.557 | 75.428 |
| For cohort genotype = CC | 2.465 | 1.788 | 3.398 |
| For cohort genotype = CT+TT | 0.150 | 0.040 | 0.568 |
| Number of active cases | 100 |  |  |

**Table 29:** Family History Risk Assessment of Cholesterol Gallstones.

|  | value | 95% confidence interval |  |
| --- | --- | --- | --- |
|  |  | lower limit | upper limit |
| Ratio of family history (with/without) | 6.689 | 2.667 | 16.776 |
| For cohort genotype = CC | 2.384 | 1.589 | 3.576 |
| For cohort genotype = CT+TT | 0.356 | 0.197 | 0.645 |
| Number of active cases | 100 |  |  |

## Discussion

Gallstone disease is currently one of the most common digestive system diseases, and the epidemiological survey shows that the prevalence of gallstones is increasing year by year. laparoscopic cholecystectomy is currently the most effective and commonly used method for the treatment of symptomatic gallstones^[25]^, but as an aggressive treatment and the harm it brings to patients due to the increasing prevalence of gallstones year by year, studying its pathogenesis has more important clinical significance. Cholesterol gallstones account for more than 70% of all gallstones, and the main exogenous risk factors include high-calorie diet, lack of exercise, obesity, pregnancy, estrogen, advanced age, diabetes, and dyslipidemia^[26]^. In recent years, researchers have made some progress in the study of the formation mechanism of cholesterol gallstones in many fields, but the underlying cause is still uncertain. At present, it is generally believed that abnormal blood lipoprotein and lipid metabolism are one of the main factors that lead to changes in the physicochemical properties of bile and its transformation to cholesterol gallstones^[4]^. At the same time, epidemiological and familial studies have found that genetic factors account for about 25% of the total risk of gallstones^[6]^, indicating that the formation of gallstones is caused by the common participation of environmental and genetic factors. With the development of molecular biology and the continuous improvement of whole genome sequencing technology, people have gradually explored the mechanism of cholesterol gallstones in the gene field.

Single Nucleotide Polymorphism (SNP) refers to the phenomenon of single nucleotide variation in the DNA sequence of different individuals in organisms. More than 90% of the genetic variations in the human genome are present as SNPs. In contrast to rare mutations, SNPs are mutations with a frequency of greater than 1% whereas rare mutations appear as mutations with a frequency of less than or equal t o 1% in a population^[27]^. To date, the total number of gallstone susceptibility genes and their polymorphic sites confirmed by genome-wide association studies (GWAS) has reached 75^[28]^. Eleven of these were associated with plasma total cholesterol levels. Among these 11 variants, some lithogenic alleles such as the ATP-binding cassette transporter G8 gene, the hepatocyte nuclear factor 4α (Heptocyte nuclear fac tor 4α, HNF4α) gene and Glucokinase regulatory protein (GCKR) gene were associated with decreased plasma total cholesterol, while other genes such as ATP-binding cassette subfamily B member 4 (ATP-binding cassette, Su B-family B, Member 4, ABCB4) gene and 7α hydroxylase (7α-hydroxylase, CYP7A1) gene were associated with increased plasma total cholesterol. The study found that the decrease of plasma TC level might be related to the increase of cholesterol liver outflow by the variants, while the increase of plasma TC might be related to the decrease of bile acid synthesis by the variants^[29–31]^.

Kruppel-like factor 14(Krüppel-like factor14, KLF14), also known as basic transcription factor binding protein 5, is widely expressed in mammalian tissues, including muscle, brain, heart, fat, and liver. The human KLF14 gene is located at 7q32.3, with a total length of 1059 bp and a molecular weight of 33124 Da. KLF14 regulates gene expression involved in many important physiological and pathological processes in organisms by binding to cis-regulatory sites rich in GT/GC in promoters or enhancers of target genes^[9]^. guo^[32]^They found in the liver of type 2 dyslipidemia mouse model that the specific deletion of KLF14 could lead to the reduction of HDL-C level in the circulation, and promoted the expression of KLF14 by regulating the production of liver ApoA-I, thereby improving HDL-C level and cholesterol efflux capacity. The results of GWAS also confirmed that the polymorphic sites of the KLF14 gene rs4731702 and rs972283 were closely related to HDL-C levels, metabolic syndrome, and atherosclerosis^[22]^. At present, the research on KLF14 gene focuses on coronary artery disease, hypercholesterolemia and type 2 diabetes^[34]^, and there are few studies on its correlation with cholesterol gallstones.

A study outside China has shown that for mice with overexpression of SR-BI gene in the liver, plasma HDL cholesterol is ingested by the liver and enters the bile through SR-BI, thus increasing the cholesterol concentration in bile [6]. By contrast, in SR-B1-deficient mice, plasma cholesterol levels are elevated, and secretion of bile cholesterol is decreased [7]. SR-B1 gene is located on chromosome 12, and 12 exons have been identified so far. Synonymous single nucleotide polymorphisms (rs5888, T[ thymine] and C[ cytosine]) located in exon 8 have been shown to be associated with the expression level and function of SR-B1[12]. The minor allele T can cause changes in the secondary structure of mRNA, resulting in decreased expression and function of SR-B1. A larger number of studies have shown that rs5888 is associated with blood lipids and coronary heart disease risk in different populations [13–18]. A study outside China has shown that the rs5888 polymorphism of SR-B1 gene is associated with higher HDL cholesterol and lower triglyceride levels in non-Asian men. We also observed an association of the rs838880-C allele of SR-B1 with increases in plasma TC[19] and HDL cholesterol [20]. At present, SNP studies on SR-B1 are mostly reported in European and American literature, and studies on Chinese population are relatively few. Due to the problems of population stratification, geographical differences and the heterogeneity of genetic background, there are certain limitations in association studies.

Therefore, foreign studies on SR-BI gene polymorphism may not be applicable to the Chinese population, and need to be verified in different populations.

The analysis in this study showed that the distribution of KLF14 gene (rs4731702, rs972283) and SR-B1 gene (rs5888, rs838880) in the case group and the control group all conformed to Hardy-Weinberg’s law of genetic balance (P > 0.05), indicating that the research subjects were randomly selected and representative of the population, and they also met the statistical requirements of genetic research. Direct sequencing was used for SNP typing. The results showed that the frequencies of genotype TT, CT AND, and CC at the polymorphic site of KLF14 gene rs4731702 in the case group were 10%, 42%, and 48%, respectively, and those in the control group were 12%, 58%, and 30%, respectively. There was a statistically significant difference in the distribution between the two groups (P < 0.05). The frequency of CC genotype in the two groups was higher than that of CT and TT (P < 0.0 5), suggesting that the mutant genotype CC might be a risk factor for chole sterol gallstones (OR = 2.154). The frequencies of C and T alleles in the case group were 69% and 31%, respectively, while those in the control group were 59% and 41%, respectively. There was significant difference in the distribution between the two groups (P < 0.05). The frequency of C allele was higher than that of T allele, suggesting that the C allele might increase the risk of cholesterol gallstones (OR = 1.547). These results indicated that the KLF14 locus rs4731702 might be associated with the development of cholesterol gallstones, and the mutant CC genotype or allele C might increase t he risk of cholesterol gallstones. We also analyzed the association of this gene locus with blood lipid indexes, gender, and ethnicity in the cholesterol gallstones group and found that the TG levels in the CC genotype of the KL F14 gene polymorphism locus rs4731702 in the case group were higher than those in the CT and TT genotypes (P < 0.05). The HDL, Apo-AI and A

po-AI/ Apo-B levels in the CC genotype were lower than those in the CT and TT genotypes (P < 0.05), suggesting a close correlation between this locus and blood lipid levels, which is also in line with the research results of GWAS^[24]^, at the same time with Chasman^[35]^In a large-scale genome-wide association study involving 17,296 European women, the findings were consistent. We concluded that the association of the KLF14 gene locus rs4731702 with gallbladder cholesterol gallstones might be related to the effect of this mutation locus on lipid metabolism.

Differences were also found between the sexes at rs4731702 in the case group (P < 0.05), compared to Small and Yang^[36]^, they found that the KLF14 gene locus rs4731702 in LDL, HDL-C, TG, the comparison of gender differences, and the effect of women is significantly higher than men. And luoxiaoqing^[37]^ The results of KLF14 gene test among the han population in qingdao, china showed that this gender difference was more significant when men carried the CC genotype and women carried the CT genotype. In the comparison between the KLF14 locus rs972283 and cholesterol gallstones, although the distribution differences of allele frequency and genotype frequency in the case group and the control group were not statistically significant (P > 0.05), there was a certain correlation between the KLF1 4 gene rs972283 and blood lipid indexes in the case group (P < 0.05), which showed that the TG and TCH levels in the GG genotype were higher than those in the GA and AA genotypes. GG genotype had lower HDL, Apo-AI and Apo-AI/ Apo-B levels than GA and AA genotype. kong^[38]^The research by et al. also showed that this site was related to the increase of TG level.

The experimental results showed that the genotype distribution of rs5888 locus of SR-B1 gene in case group and control group conformed to Hardy-Weinberg’s law of genetic balance (P > 0.05), and three genotypes (CC, CT and TT) could be detected in both groups. With the sample size n≥40 and all T≥5, Pearson Chi-Square test was selected and the results showed that the gene frequencies of CC, CT and TT in the case group were 66%, 29% and 5%, respectively, while those of genotype in the control group were 46%, 45% and 9%, respectively. The difference in the distribution between the two groups was statistically significant (P < 0.05). The CC genotype at rs5888 of SR-B1 gene in the case group was significantly higher than that of CT+TT genotype (P < 0.05), suggesting that the mutant CC genotype might be a risk factor for cholesterol gallstones (OR = 2.279). The frequencies of C and T alleles in the case group were 80% and 20%, respectively, while those in the control group were 68% and 32%, respectively. There was a significant difference in the distribution between the two groups (P < 0.05). The frequency of C allele was higher than that of T allele, suggesting that the C allele might increase the risk of cholesterol gallstones (OR = 1.898). However, there was no significant difference between the case group and the control group in the analysis of S R-B1 gene rs5888 and gender, ethnicity (P > 0.05). Through the analysis of rs5888 and related lipid indicators, we found that in the case group, the gene frequency of people with TT type gene was significantly lower than that of the control group, the HDL-C of people with TT type gene was significantly higher than those of CC and CT type, while the contents of TG and TCH were significantly lower than those of CC and CT type (P < 0.05). There was no statistically significant difference in LDL, Apo-AI, Apo-B and Apo-AI/ Apo-B (*P* > 0.05). Therefore, we can infer that rs5888 of SR-B1 gene may have a direct relationship with the occurrence of gallbladder cholesterol gallstones, an d genotype CC type may be the risk genotype for the occurrence of gallbladder cholesterol gallstones, which is consistent with the results of Ruchao et al. [39].

Three genotypes (TC, TT and CC) were detected through genotyping of the SR-B1 gene at rs838880 in the case group and the control group. With the sample size n≥40 and all T≥5, Pearson Chi-Square test was selected and the results showed that the gene frequencies of TC, TT and CC in the case group were 48%, 49% and 3%, respectively, while the genotypic frequencies in the control group were 33%, 36% and 31%, respectively (*P* < 0.0 5). The number of CC genotype at rs838880 of SR-B1 gene in case group was significantly lower than that in control group, suggesting that genotype CC at rs838880 of SR-B1 gene might be the protective genotype for cholesterol gallstones (OR=0.116). The frequencies of C and T alleles in the case group were 27% and 73%, respectively, and those of the control group were 47% and 53%, respectively. The difference in distribution between the two groups was statistically significant (*P* < 0.05). The frequency of T allele was higher than that of C allele, suggesting that the C allele might reduce the risk of gallbladder cholesterol gallstones (OR = 0.409). At the same time, the HDL of CC genotype was significantly higher than that of TC and TT genotype in the case group, and the difference was statistically significant (*P* < 0.05), which also indicated that the C allele might be related to the increase of HDL-C in blood. In the research by Victor Lazarenko et al. [40], t hey believed that there was a certain correlation between the C allele and the increased levels of TCH and LDL-C, which was related to the increase of TCH and HDL-C in plasma. The author believes that the results of this experiment are only partially consistent with those of Victor Lazarenko et al., which may be due to differences in population selection, geographical distribution, national customs and culture, etc.

The current research has found that many diseases are caused by multi-gene interaction rather than single gene.^[41–42]^. The same is true of gallstones. The interaction of multiple genes and their polymorphic sites leads to the occurrence of gallstones^[43]^, at the same time also cannot ignore the importance of environmental factors. In addition, researchers have found for a long time that different people have different susceptibilities to the same disease. due to the existence of gene polymorphism, different individuals with the same disease also have differences in clinical manifestations, which indicates that genes have produced differences in the mutation process^[44]^. Similarly, polymorphism of drug metabolism-related genes can affect drug efficacy by affecting the metabolic process of drugs in the human body, resulting in differences in the efficacy of drugs in different individuals^[45]^. Therefore, in this study, the association between locus rs4731702 and locus rs972283 of KLF14 in the case group was analyzed, and the results showed that although there might be no association between locus rs972283 of KLF14 and cholesterol gallstones, the above two loci might have a certain correlation in the pathogenesis of cholesterol gallstones. However, through studies on the independence of the two genes and the association between rs5888 and rs838 880 of SR-B1 in the case group, we believe that the above two genetic loci may play their respective roles in the pathogenesis of cholesterol gallstones, but they do not have a certain association. Then we analyzed the association between the SR-B1 gene locus rs5888 and the KLF14 gene locus rs4731 702 in the case group, and we thought that the above two gene loci might not only play their respective roles in the pathogenesis of cholesterol gallstones, but also have a certain correlation.

At the same time, we also need to consider the differences in susceptibility brought by environmental, regional and ethnic customs and other factors. In this study, by means of case inquiry and questionnaire, the BMI, family history of gallbladder cholesterol stones, smoking, drinking, drinking tea, and exercise habits of the population in the case group were investigated and counted. We found that the elevated BMI and family history of gallbladder cholesterol stones were the risk factors for the occurrence of gallbladder cholesterol stones.

Exercise habit is the protective factor of cholesterol gallstones. Immediately after that, we conducted research on the interaction between genes and environmental factors. The research showed that patients with genotype (KLF14 gene rs4731702) of CC type with BMI > 26 were more likely to suffer from cholesterol gallstones (OR=16.379) than patients with BMI < 26. Patients with a family history of gallstones of genotype (KLF14 rs4731702)CC had a 5.689-fold increased risk (OR=6.689) of developing CC compared with patients without a family history of gallstones of genotype (KLF14 RS 473702) (P < 0.05). Therefore, we can conclude that the interaction between genetic factors and environmental factors promotes the occurrence of cholesterol gallstones. This result is consistent with that of Koebnick C ^[46]^. However, our result is not completely consistent with that of Wang et al. Through discussion and research, we analyze the differences in conclusions and believe that the reasons for such differences can be attributed to the differences in national customs and cultures, the differences in daily dietary structure and the differences in national regions.

First mouse gallstone susceptibility gene since 1995^[47]^ The first A TP-binding cassette transporter G8 (ABC G8) gene of human lithogenic gene was found.^[48]^ Since its inception, human beings have been developing on the road to explore the genetic mechanism of cholesterol gallstones. To date, GWAS has confirmed that the total number of gall bladder stone susceptibility genes and their polymorphic sites has reached 75. An in-depth study of the gallbladder cholesterol stone-related susceptibility genes and their polymorphisms is crucial for further studies on the genetic mechanisms of cholesterol stone formation, as well as for early screening and personalized prevention. However, due to the fact that gene polymorphisms are affected by geographical and ethnic differences, the results of the studies vary significantly, and the occurrence and development of gallbladder cholesterol gallstones are related to multiple genes, there is still a lack of adequate understanding of the correlation between each gene and the determination of the dominant gene, as well as the short collection time and limited number of samples included in this study, which preclude more accurate and detailed statistical analysis. Therefore, in the future, we need to add multiple environmental factors and genes, and gene-gene correlation analysis on the basis of expanded sample size, and determine the relationship between gene polymorphism and cholesterol gallstones through the increasingly developed whole genome sequencing technology and large-scale, multi-center clinical trials.

In conclusion, cholesterol gallstones is a common digestive system disease that occurs under the joint participation of genetic and environmental factors. The important role of susceptible genes and their polymorphisms in the occurrence and development of cholesterol gallstones has been paid attention by scholars both in China and abroad. This study has revealed that the polymorphisms of KLF14 gene and S R-B1 gene may be associated with cholesterol gallstones, and there are certain interactions between genes and genes as well as between genes and the environment, which not only provide new ideas for further research on the genetic mechanism of cholesterol gallstones, but also may provide new theoretical basis for early diagnosis and personalized prevention.

## Conclusion

1. The SNP at rs4731702 of the KLF14 gene may be related to the susceptibility to cholesterol gallstones, and the carrying of allele C may increase the risk of cholesterol gallstones.
2. Genotype CC of the SR-B1 gene rs5888 may be the risk genotype for the development of cholesterol gallstones.
3. Genotype CC of the SR-B1 gene rs838880 may be the protective genotype for cholesterol gallstones, and the C allele may be associated with increased HDL-C in the blood.
4. Single nucleotide polymorphisms in the KLF14 and SR-B1 genes were significantly associated with dyslipidemia and lipoprotein metabolism.
5. Patients with cholesterol gallstones have varying degrees of abnormal lipid and lipoprotein metabolism in blood.
6. The increased BMI and family history of cholesterol gallstones we re the risk factors for cholesterol gallstones. Exercise habit is the protective factor of cholesterol gallstones.
7. The interaction between genetic factors and environmental factors promotes the occurrence of cholesterol gallstones.

## Data Availability

All relevant data are within the manuscript and its Supporting Information files.

